# Black Women’s Lived Experiences of Depression and Related Barriers and Facilitators to Utilising Healthcare Services: A Systematic Review and Qualitative Evidence Synthesis Co-produced with Experts by Lived Experiences

**DOI:** 10.1101/2024.09.02.24311928

**Authors:** Anna-Theresa Jieman, Farida Soliman, Keisha York, Kamaldeep Bhui, Juliana Onwumere, Sanisha Wynter, Faith Amasowomwan, Sandra Johnson, Janelle M. Jones

## Abstract

Depression among Black women is a significant public health concern. However, our understanding of their unique experiences and the barriers and facilitators to utilising healthcare services remains limited. To address these issues, we conducted a qualitative evidence synthesis in collaboration with experts by lived experiences. We searched seven databases (ASSIA, MEDLINE, APA PsycInfo, Sociological Abstracts, CINAHL, AMED and EMBASE) from inception to 9^th^ September 2021 and updated to 29^th^ March 2024 with an English language restriction. Study quality and confidence in findings were assessed using the Critical Appraisal Skills Programme (CASP) and Confidence in the Evidence from Reviews of Qualitative Research (GRADE-CERQual) approach. Of 15025 papers screened, 45 were eligible for inclusion. Data were analysed using thematic analysis. Women reported depression stemming from racial and gender-related stressors, social isolation, and a loss of faith; moreover, the ‘Strong Black Woman’ schema masked depression symptoms. Mistrust of healthcare providers, stigma, religious coping, and pressure to conform to the Strong Black Woman schema hindered healthcare service utilisation. The rapport between women and their healthcare providers, endorsement from faith leaders, and points of crisis enabled service utilisation. Lived experience experts provided reflections and recommendations for practice.

**Highlights:**

- Recognition of depression may be hampered by schemas connected to Black women’s identity.
- Trust between Black women experiencing depression and clinicians is essential for effective care.
- Training which incorporates antiracist principles is needed for competence in discussing issues surrounding race and gender.
- (Re-)consideration of diagnostic criteria to acknowledge differential presentation and the development of culturally adapted treatments are warranted.
- Co-producing research with experts by lived experience ensures it is more impactful.

## Introduction

Depression is a major global mental health concern, affecting approximately 3.8% of the world’s population[1, 2]. Characterised by persistent feelings of sadness, loss of interest or pleasure, poor concentration, and fatigue, depression has been associated with considerable losses in health and functioning[2, 3] and death by suicide[4–6]. Depression disproportionately affects women worldwide (prevalence: 6% for women versus 4% for men)[1]. The difference in prevalence has been attributed to various factors, including gender-based violence[7]. In the United Kingdom (UK) and the United States of America (USA), there are racial and ethnic disparities and inequalities in prevalence and seeking and receiving treatment for depression. In the UK, Black women have the highest rates of depression[8]. In the USA, the lifetime prevalence of depression was highest among White Americans (17.9%) compared with Black Caribbeans (12.9%) and African Americans (10.4%)[9]. Among African Americans, women had almost twice the rate of depression compared with men. Although White Americans had greater prevalence compared with Black individuals, the study concluded that depression was more chronic among Black Caribbeans and African Americans, as the measure of persistence over 12 months was higher for Black Caribbeans (56.5%) and African Americans (56%) than White Americans (38.6%)[9]. Black individuals may have an increased risk and chronicity of depression due to experiences of racism and discrimination[10–12]. Therefore, a review focused on Black women’s experiences is crucial to understanding how racism, gender-based discrimination and other factors might be contributing to increased risk and chronicity.

Although Black women are susceptible to experiencing greater prevalence and more chronic depression in the UK and the USA, they are less likely to utilise healthcare services. Research suggests that only 45% of African Americans and 24.3% of Black Caribbeans sought treatment[9]. However, they were more likely to rate their depression as more severe and disabling compared with their White American counterparts[9]. Furthermore, recent studies show that Black individuals are underrepresented in non-emergent healthcare utilisation in the USA[13, 14], including mental health services[15], but overrepresented in emergency services[14]. These trends remain among individuals who use Medicaid, where the majority of the enrolees are racially minoritised individuals[14]. Some of the barriers documented are related to stigma, cost, logistics, discrimination, and life difficulties[16]; lack of access to culturally competent healthcare providers[17]; mistrust of healthcare providers (HCPs); and the notion that seeking treatment for mental illness is a sign of weakness which should be resolved within the family or through religious coping[18]. In the UK, most individuals have access to free healthcare via the National Health Service (NHS), which allows them to also self-refer to specialist mental health services such as the NHS Talking Therapies for Anxiety and Depression. However, Black individuals continue to be underrepresented in these services[19–21]. A study investigating referral pathways (e.g., self-referral and via general practitioner)[22] found that once Black individuals were referred, they were less likely to receive an assessment. Furthermore, among those who were referred, Black individuals were less likely to receive treatment compared with White individuals[22]. Hence, greater insight into barriers and facilitators to service utilisation is needed.

Given these inequalities and disparities, we sought to conduct a qualitative evidence synthesis (QES) to gain insight into the nature of how Black women experience depression and how and why it influences diagnosis and barriers and facilitators to accessing and utilising services. Qualitative methods are ideal for critically investigating the under-researched, complex, and multifaceted structural root causes of illness, such as racism and power[23, 24]. They play a significant role in understanding health inequalities and disparities and their determinants because they can illuminate the underlying social, cultural, and political factors across all levels of the social and ecological framework in a manner representative of individual experiences and circumstances[25, 26]. Moreover, qualitative methods have the potential to generate new theories of change between constructs[25, 26]. Therefore, like others[27], we aimed to develop a logic model linking specific experiences to barriers and facilitators to utilising healthcare services to mitigate the health inequalities and disparities we have identified.

This QES explored intersecting forms of oppression by focusing on the Strong Black Woman Schema (SBWS), a set of beliefs and behaviours that are ‘expected’ of Black women. According to this schema, Black women should exhibit strength through independence, hard work, emotional suppression, caregiving, and self-sacrifice[28]. Research from the USA indicates that endorsement of the SBWS is associated with depression and anxiety[29, 30] and perceptions that one lacks emotional support[31]. Although research on the SBWS has been conducted in the USA, Black feminist theorists[32] suggest that all Black women in the African diaspora share a collective history that includes slavery, colonialism, involuntary migration, and face double jeopardy oppression due to their race-gender intersected identity[33]. These historical oppressions and experiences seem to influence present issues, given the prevalence of sexist, racist, and classist principles that persist within countries where Black women are racially minoritised individuals. Thus, Black women may share ideas about what it means to be Black and a woman, which could considerably influence how they construct and experience depression and approach help-seeking. Therefore, we contend that the internalisation of societal beliefs about Black womanhood may lead Black women to minimise their difficulties, ultimately influencing the experiences of depression and creating barriers to accessing help.

## Objectives

This review aimed to identify, appraise, and synthesise qualitative research on Black women’s experiences of depression and barriers and facilitators to accessing help.

The following research questions guided this objective:

1a. How do Black women experience depression, and how does this influence related help-seeking behaviours?
1b. In what ways might race and/or gender and the Strong Black Woman Schema influence Black women’s experiences of depression and related help-seeking behaviours?

## Methods

This QES followed the updated Preferred Reporting Items or Systematic Reviews and Meta-Analyses statement guidelines (Appendix A)[34] and reporting guidelines from Enhancing Transparency in Reporting the Synthesis of Qualitative Research (ENTREQ) (Appendix B)[35]. The protocol for this review was registered on PROSPERO: <u>CRD42022304571</u>.

### Criteria for study inclusion

Participants were Black women aged 18+, including those of African and Caribbean descent and dual heritage. If we could not identify which excerpts belonged to Black women in mixed participants studies, authors were contacted to clarify. Exclusions applied to studies that did not include or focus on Black women.

We included qualitative studies investigating experiences of depression or low mood (as a primary concern) and related help-seeking behaviours. No formal diagnosis of depression was required, acknowledging previous research indicating lower rates of help-seeking in this population[9, 19–21]. Help-seeking included various coping strategies, both adaptive (e.g., utilising healthcare services, religious coping) and maladaptive (e.g., excessive alcohol consumption, self-harm). Studies that focused on postpartum depression were excluded as postpartum depression has distinct psychopathology and pathophysiology[36].

The included studies were limited to those conducted in high-income countries (as defined by the World Bank) where Black women are considered a racially minoritised group. Settings encompassed healthcare (primary, secondary, and tertiary) and community environments. Exclusions applied to studies conducted in middle, low, and high-income countries where Black women are not considered a racially minoritised group (e.g., Barbados).

Eligible studies were primary qualitative studies published in academic journals or PhD theses, including mixed methods with available qualitative data excerpts. Exclusions included abstracts, conference proceedings, case reports, book chapters, opinion pieces, vignettes, editorials, and non-English publications.

### Search methods

AJ conducted the searches. The databases searched were ASSIA, MEDLINE, APA PsycInfo, Sociological Abstracts, CINAHL, AMED, and EMBASE from inception to 9^th^ September 2021 and updated to 29th March 2024. Boolean searching with free-text and MeSH terms were employed (e.g., depression OR “low mood” AND Black OR “African American” AND woman OR female). The search strategies for a comprehensive search (Appendix C) underwent iterative refinement, considering relevance. Reference lists of included studies and critical systematic reviews were also reviewed to identify potentially relevant studies. Finally, we hand-searched relevant journals, consulted leaders of community organisations in the UK, and contacted authors globally if we could not identify which data belonged to Black women in mixed population qualitative studies.

### Study selection

Studies were deduplicated in Mendeley and then hierarchically screened on title and abstract and full text in EPPI Reviewer. Twenty per cent of the papers were double-screened on the title and abstract, and all papers were double-screened in full text by AJ and FS. Based on best-practice guidelines[37], AJ and FS piloted all screening tools to ensure consistency. All disagreements were resolved through discussion and, if necessary, consultation with a third author (JJ, KB or JO).

### Assessment of methodological limitations

The Critical Appraisal Skills Programme (CASP)[38] tool for qualitative studies was used for quality appraisal. We did not exclude low-quality or inadequately reported studies, as previous researchers found that including them did not contradict the themes developed from higher-quality studies[39–41] but did impact the transferability of findings[40]. Therefore, following guidance, the results informed the analysis sequence (i.e., well-reported studies were coded first) rather than exclusion. AJ and FS independently applied the CASP tool, with discrepancies resolved through discussion.

### Data extraction

Data extraction occurred in two stages. First, FS and AJ independently extracted contextual information concurrently with quality appraisal. If duplicate data from a project with multiple papers were found, the information provided in each paper was extracted. However, excerpts were not duplicated for the analysis. Second, AJ and KY independently extracted quotes from participants.

### Data management, analysis, and synthesis

Our approach to Patient and Public Involvement and Engagement (PPIE) occurred during the research question development, analysis, interpretation, and writing stages, following a similar method used by other researchers who have co-produced and written systematic reviews with experts by lived experiences[42–44]. Our PPIE group included a clinical psychologist (JO) and experts with lived experiences (SW, FA, and SJ) who provided reflections (included verbatim in the results), actively participated in the manuscript development and were invited to be co-authors. The PPIE group were remunerated according to guidelines for participatory research[45].

The thematic analysis broadly followed the approach outlined by Thomas and Harden[46]. AJ and KY used EPPI Reviewer, which allows for line-by-line coding. Initial codes were presented to the research team for feedback, leading to adjustments for accessibility. Then, AJ and KY coded all the included papers, developing descriptive themes. The initial coding and development of descriptive themes was deductive as AJ and KY coded data as an experience, barrier, or facilitator based on previous research. Subsequently, it was inductively revised as new codes and themes emerged. AJ aligned descriptive themes to the research questions to create analytic themes, forming a narrative. Team discussions and theory identification informed adjustments until a consensus was reached. AJ associated each experience with a corresponding barrier and facilitator to determine how each experience of depression may influence help-seeking behaviours, leading to the development of a logic model. The findings were discussed with the PPIE group for reflections and recommendations. The whole review team (led by AJ and KY) were involved in the analysis, synthesis and interpretation, fostering reflexivity and rigour. The PPIE process, analysis, and synthesis are summarised in Figure 1.

**Figure 1:**
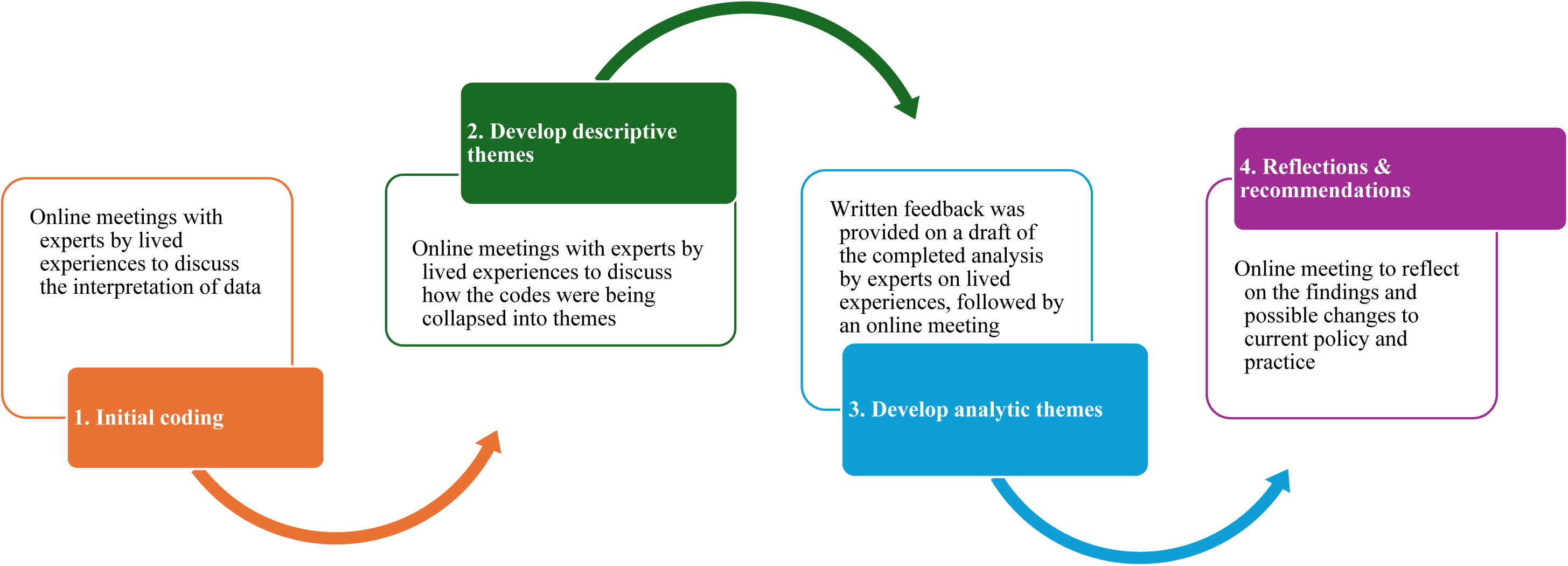
Stages of analysis and patient and public involvement and engagement.

## Review team reflexivity

The review team comprised individuals with diverse professional backgrounds and expertise, including clinical researchers, researchers at different levels, and experts with lived experiences. They all identified as racially minoritised individuals (seven were Black women, one South Asian man, and one assigned female at birth North African).

The team engaged in reflexive discussions throughout the analysis to enhance transparency, rigour, and awareness of potential biases. This led to a more nuanced understanding of symptoms, experiences of depression, and stereotypes associated with Black women. For instance, AJ was initially sceptical about reporting that Black women experience anger as a symptom, in fear of reinforcing that Black women fit the ‘Angry Black woman’ stereotype. However, she reported anger as it is a typical symptom, and she realised that not reporting experiences of depression that might be associated with negative connotations was not reinforcing stereotypes. Instead, it showed that depression has various presentations across individuals due to different explanatory models of illnesses. Not considering these explanatory models leaves Black women disadvantaged and reinforces disparities and inequalities.

### Assessing confidence in the findings

AJ and FS used the GRADE-CERQual (Confidence in the Evidence from Reviews of Qualitative Research) approach to assess confidence in each finding[47]. After independently assessing each of the four components (relevance[48], methodological limitations[49], coherence[50] and adequacy[51]) of the GRADE-CERQual, we assessed the overall confidence[52]. All findings were initially judged as high confidence and degraded if there were essential concerns regarding any GRADE-CERQual components. The final assessment was based on consensus among the review authors.

## Results

### Results of the search

Our search retrieved 24836 papers with 9811 duplicates. After the screening (see Figure 2), 45 papers met the inclusion criteria. The papers were published between 1994 and 2023 and conducted in the USA (n=36), the UK (n=5), and Canada (n=4).

**Figure 2:**
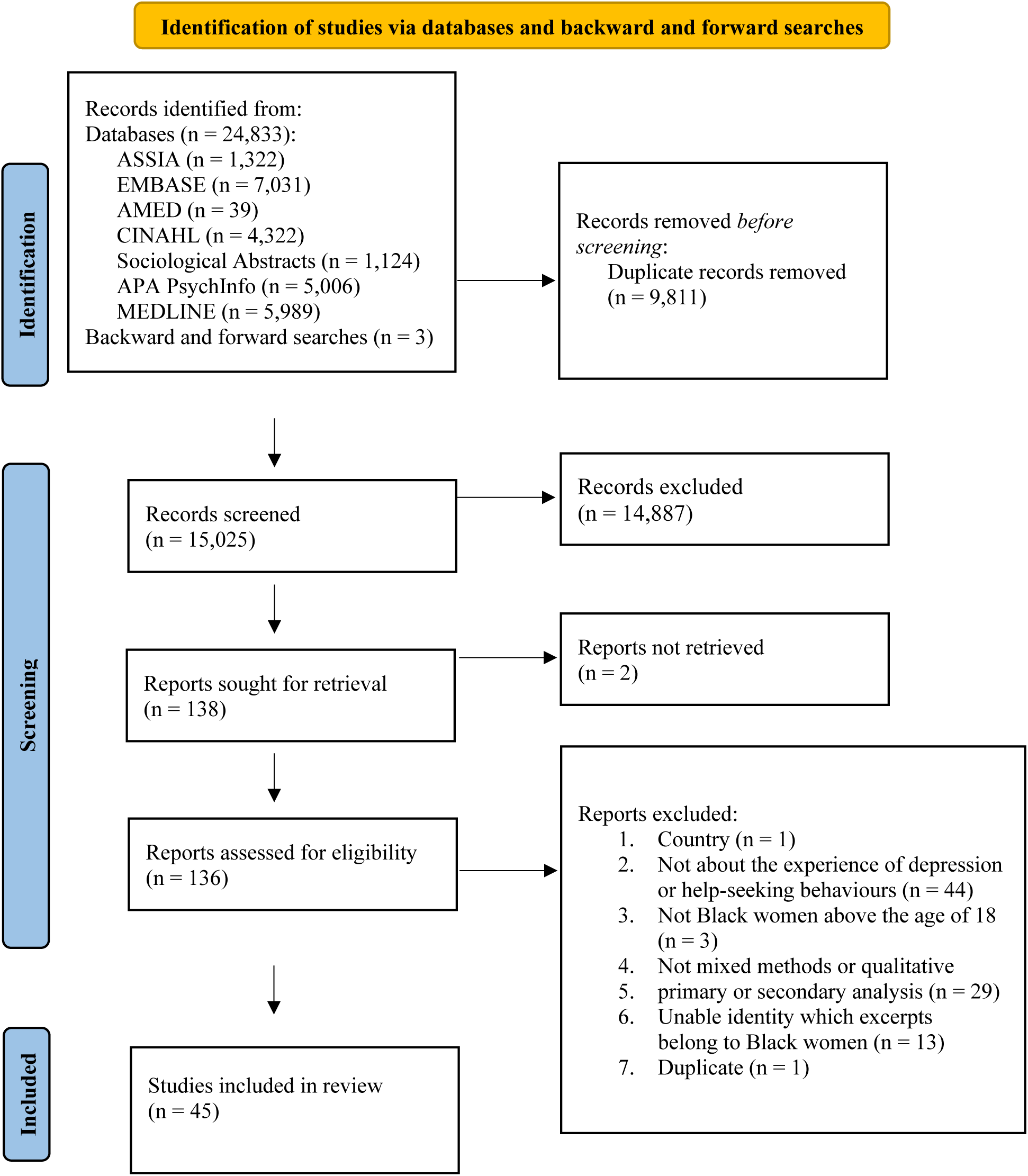
PRISMA flow diagram.

### Study characteristics

All studies used qualitative or mixed-methods study designs. Common data collection methods were in-depth interviews and focus groups, and the most common method of data analysis was thematic analysis. Sample sizes ranged from 5 to 63. The age ranged from 18 to 78 years. Thirty-one papers measured levels of depression with either a validated questionnaire (e.g., Patient Health Questionnaire 9) or asked participants to self-disclose previous experiences of depression. Nine papers did not measure experiences of depression. Five papers did not state whether they measured experiences of depression. Ten papers reported that participants had been treated for depression. Twenty-seven papers did not state whether participants had been treated for depression, and eight papers reported that participants had not been treated for depression (see Appendix D for included study characteristics).

### Methodological limitations of the studies

Concerning the methodological limitations of the papers included, researcher reflexivity was not reported in seven papers. All but one study described a clear research aim, methodology, recruitment strategy, analysis approach, findings, and contributions to existing knowledge. It was unclear how the authors selected their research design in three papers. In six papers, it was unclear whether the authors had sought ethical approval (see Appendix E for the CASP assessments).

### Confidence in the review findings

We assessed ten themes as high confidence and one as low confidence. Our concerns for the theme, which we scored as low confidence, were associated with moderate concerns for relevance, as two of the three studies contributing to the theme recruited a mixed sample, not only Black women. Table 1 reports the GRADE-CERQual assessments (see Appendix F for a detailed account of each element).

**Table 1:**
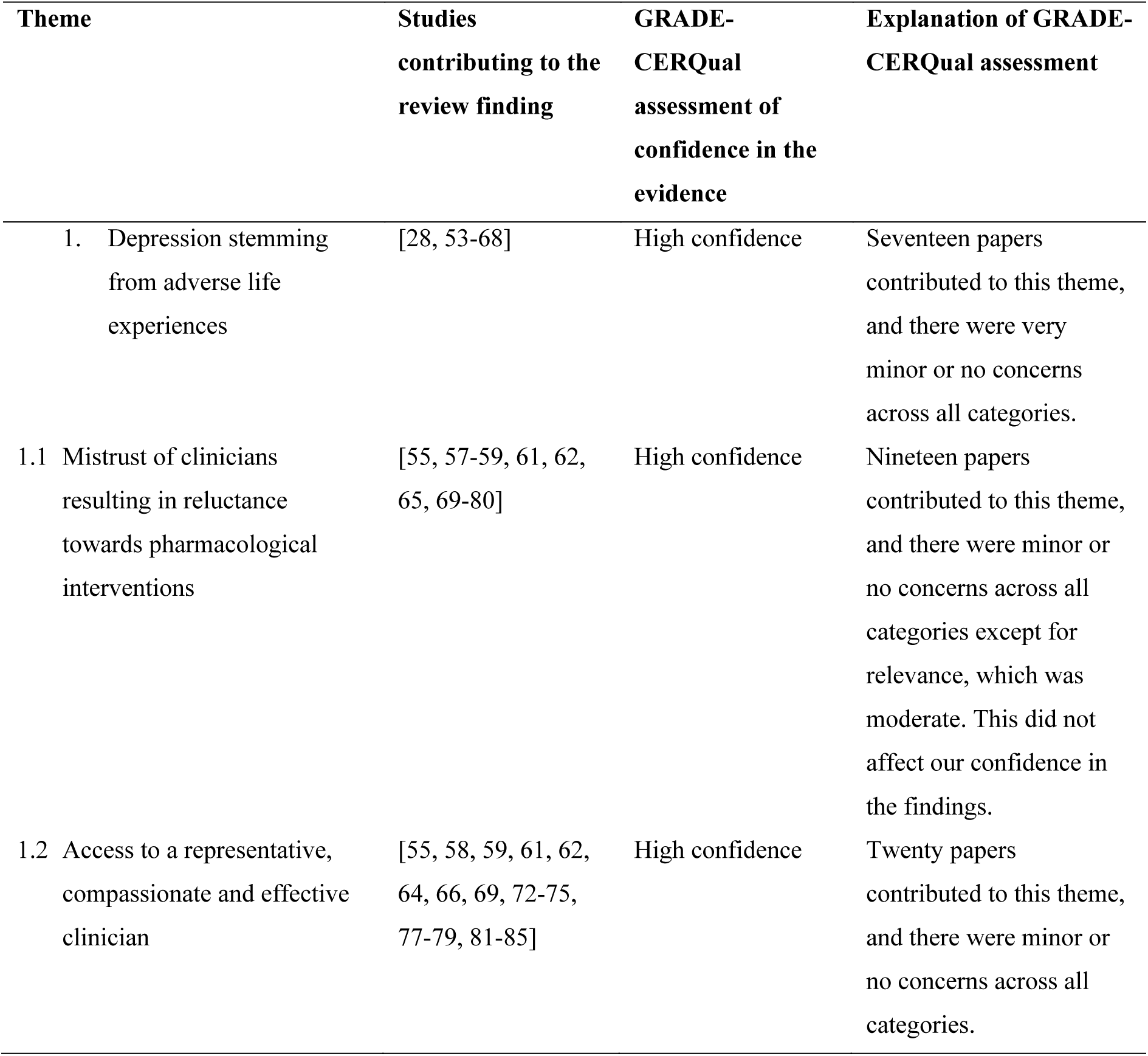
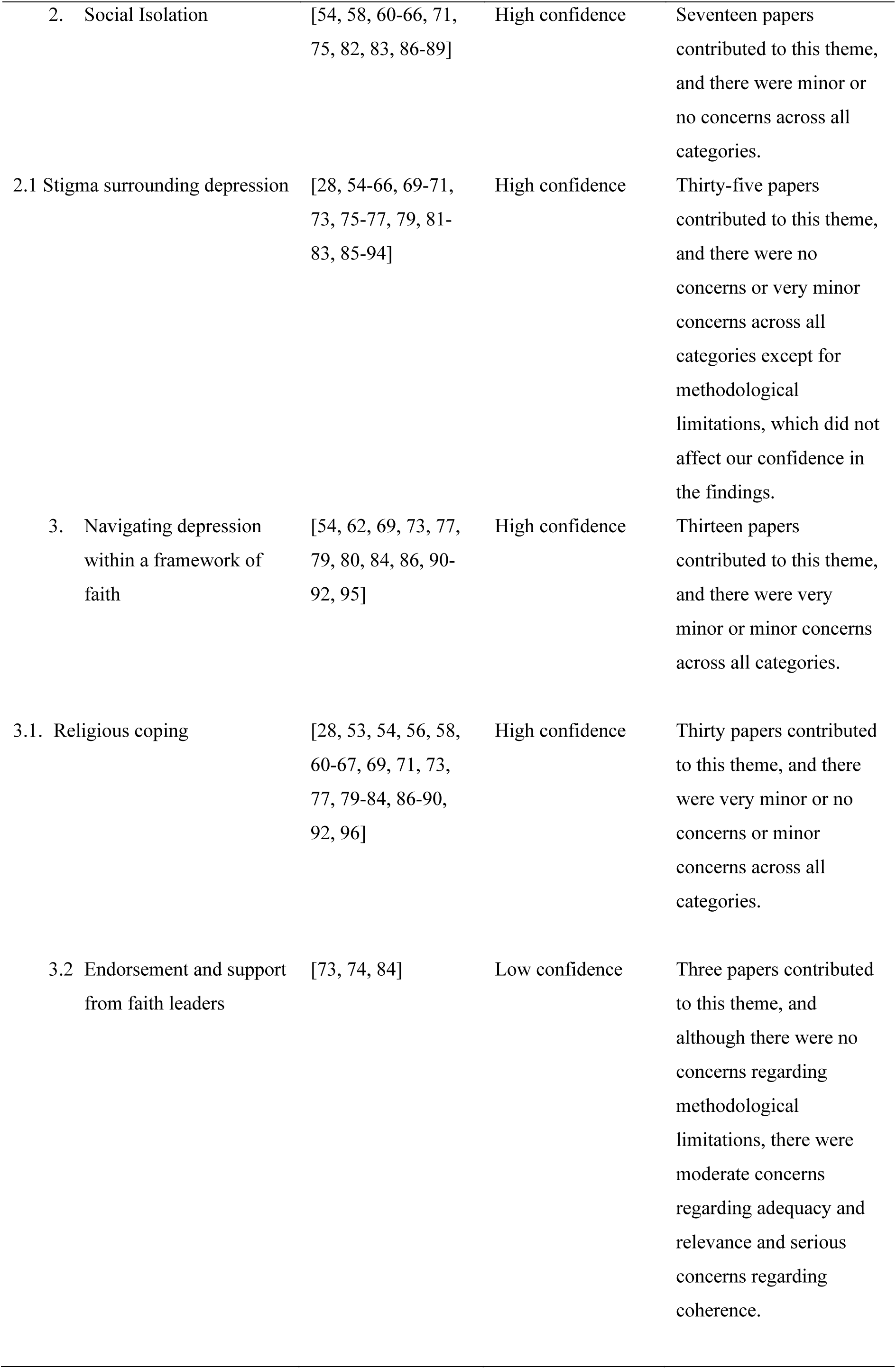
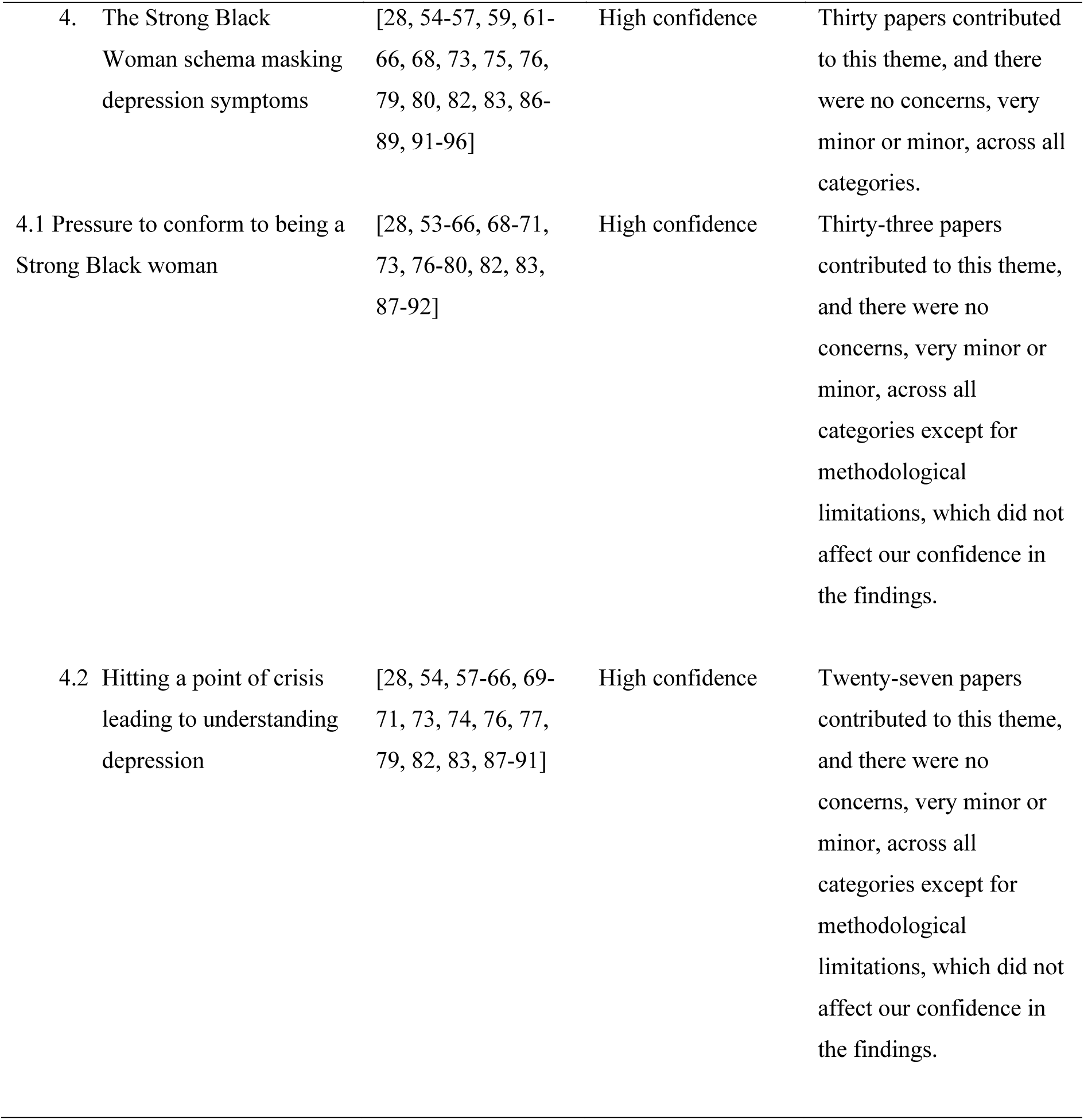
GRADE-CERQual assessment results.

### Review findings

#### 1. Experience: Depression stemming from adverse life experiences

Black women reported experiences of depression due to adverse life experiences[28, 53–66] and trauma[28, 53–66]. Although the type of adverse life experience was not always disclosed[58] when mentioned, adversities included poverty[54, 62, 64], racial trauma[56, 57, 65–67], abuse (domestic;[56, 58, 60, 63] childhood;[56, 58, 63, 65] sexual;[62, 65]), the pressure of being a single mother[68], and neglect from Black men, who were their romantic partners[28, 53, 57, 60, 61, 64]. Neglect from their partners was associated with women feeling unsupported as they were left to bear the burden of familial responsibility, which was connected to depression:

> *“There were White families, Chinese families… Indian families… and then there were Black women and children. Now I ask you… where are the black men… and why aren’t they there with their kids, anyway?”* [60]

> *“The part about neglect from Black men triggers me. I feel that when it comes to relationships, Black relationships, there is an experience that we have as Black women that many races probably cannot relate to. It’s so true that a lot of Black women have so much trauma based on their relationships*.” FA

Racial trauma was cumulative, leading to racial battle fatigue (i.e., “the physiological and psychological strain exacted on racially marginalised groups and the amount of energy lost dedicated to coping with racial microaggressions and racism”[97]) and, in some cases, a mental health crisis. Mental health crises were prevalent in the workplace, where women felt they were not valued regardless of their dedication[65, 67, 68]. They were constantly overlooked for promotions, which was associated with feeling as though their work roles were tokenistic (i.e., superficial roles used to meet the diversity needs of an institution without genuine value or acknowledgement from colleagues):

> *“…I don’t like to say it, but I think it’s that they’d got ‘The Black Person’ and as long as they got ‘The Black Person’ that’s all that matters … Being in the workforce as a Black woman, it’s like you have to prove yourself over and over*”[65].

> *“There is a fear that you will not be believed because even though many understand Black people’s experiences, the wider population think that could never happen. Unfortunately, some people are still clouded, including clinicians. So, it is the fear of giving up because you’re tired of fighting. But we must fight*.” SJ

> *“My fears concerning these results are that they are going to be overlooked, underplayed, or aligned with other social indicators because of a lack of acceptance of the impact of trauma. They might take away from some of the core issues these experiences try to bring to life by making excuses or not listening to the quotes*.*”* SW

##### 1.1 Barrier to utilising healthcare services: Mistrust of clinicians resulting in reluctance towards pharmacological interventions

Women did not trust healthcare providers[55, 58, 59, 62, 69–74] because they feared they would be treated poorly[61, 69, 71, 73, 75], that their ability to care for themselves and their loved ones would be questioned[55], there would be a breach of confidentiality and that reports of depression on their medical records would impact their careers[70, 71], and experiencing additional racial trauma from White clinicians[59, 71]:

> *“[Health care workers] are just more cold, like emotionally something happened to you that’s traumatic, they’re very cold. But if somebody that’s White come in with the same case, they’re “Oh, what’s the matter,” taking their time and you know, getting very involved personally with that person.”* [59]

There was a reluctance towards pharmacological interventions for various reasons[55, 57, 59, 62, 65, 73, 75, 76]. Black women were concerned they would be overmedicated and become addicted[55, 59, 62], they were being experimented on[73], that clinicians were only prescribing medication to fund pharmaceutical companies[59], and that taking medication made them weak[65, 76]:

> *“I did not feel like there was something evil in the way that doctors or therapists were trying to push me on medication. However, I thought it was an over-prescribing of medication instead of talking therapy, which suited my needs. Something that I believe speaks powerfully to the barriers to Black women accessing certain health service*s.” SW

> *“White clinicians tend to push medication more often and send you on your way instead of referring you to talk therapy. However, I have also had quite bad experiences with Black clinicians; they are anti-medication for mental ill health. Therefore, you have White clinicians who are very pro-medication and then Black clinicians who are the complete opposite. I feel there is a lack of a middle ground.”* FA

> *“I am on medication now, but I have not been on medication for a few years; I did try to avoid it due to fear and the stigma. I have seen a difference in me on the medication. But do I really need it? I do not know. I don’t think I do. But to meet specific criteria to be part of this healthcare system, I am taking this medication*.” SJ

Although women were reluctant to take medication, they were open to exploring talking therapies[57–59, 61, 62, 73, 75, 77]. Talking therapies allowed women to become acquainted with their psychotherapists and gradually build relationships with them. Once a trusting therapeutic relationship was established, some psychotherapists convinced women to take medication alongside their talking therapy[73]. However, some women could not build a rapport with their clinicians due to a lack of continuity of care[61, 78]; therefore, they terminated treatment[55, 71–73, 75, 79, 80]. Women who had consistent access to healthcare services reported that if they still did not trust their psychotherapist after a few sessions, they terminated treatment prematurely[73, 75]. Thus, the rapport between the healthcare provider and patient played a significant role in whether women continued to utilise mental health services:

> *“My therapist helped me through a really bad time. They helped me decide to use some medicine to help me, too. It did help, but when I felt better, I stopped that stuff because taking it too long is no good for you. When I need to, I still see my therapist.”* [73]

##### 1.2 Facilitator to utilising healthcare services: Access to a representative, compassionate and effective clinician

Black women perceived clinicians who did not have similar demographics or lived experiences as incompetent because they felt clinicians could not relate to their experiences[55, 58, 61, 62, 64, 73, 79, 81]. Some women who had experienced racial trauma wanted a practitioner of the same race[55, 59, 61, 64, 69, 75] because they wanted someone who they thought represented their experiences[55, 58, 59, 61, 62, 64, 69, 75, 77, 79, 85]:

> “*I think the way the care system set up in England and Wales; within that, you should be able to find an advocate who can represent you because a lot of what’s going on, we are misrepresented. I have very strong views of that; A lot of things are going on in our lives as Black people in this country, we are misrepresented.”* [69]

For other women, race did not matter. They wanted a psychotherapist who had lived experiences of depression[58, 59], was caring [72, 74, 75, 81], or offered them effective treatment[58, 59, 64, 66, 72, 73, 75, 78, 79, 81–84]:

> *“I see a therapist who has been good to me. They help me see what’s happening in my life so I can try and make sense of it all.”* [73]

> *“I have had quite negative experiences with a Black clinician where they have told me that I do not have any mental health issues. They have said that “if I’m diagnosed with something, then it will affect my career negatively”. So, I agree with needing to have somebody who relates. Somebody to tell you that you are not crazy and that what you’re experiencing is normal*.” FA

> *“I have always advocated for an understanding psychotherapist instead of someone who was of the same race as me. Previously, the Black psychotherapist I was talking to tried to encourage me not to let other Black people down with my depressive feelings. They wanted to motivate me in a way that I found oppressive instead of supportive. If I had a kind or culturally understanding practitioner, I would not have felt like that*.” SW

> *“I was referred to an organisation where all their therapists are Black. I had a Black male therapist, and unfortunately, my experience was not great. I was very open and transparent about how I was feeling and thinking. I suppose the therapist must make you reflect, but he made me reflect in quite a negative way, so I did not want to go back.”* SJ

#### 2. Experience: Social Isolation

Some women experienced depression as bouts of wanting to be left alone[58, 60–62, 64–66, 83, 86, 87, 89] and feeling lonely/neglected[54, 63, 64, 71, 75, 82, 86, 88]. Women wanted to be alone for multiple reasons: “being in a mood” and feeling the need to withdraw from social interactions[87], needing space to recalibrate because the presence of others triggered negative feelings[66, 89], and feeling that they were no longer welcome in their communities after disclosing struggles with depression[86]:

> *“Well, I would say after that experience I distanced myself from the church. And not feeling welcomed or supported, or loved, I distanced myself and haven’t, haven’t been the same since.”* [86]

> *“I have noticed depression among my friends who are Black women does present as hostility, so it reminds me of that. When they are at their worst, they push people away more. It is almost as though the more support you want to give them, the further away they push back and want to keep to themselves.”* FA

> *“This experience of social isolation reminds me of my current life and how I always go about life. I think being isolated protects the Black woman that I am. It provides a sense of safety and allows me to control who is around me, what’s around me, and what energies impact my day-to-day experience.”* SW

> *“When I was unwell, I wanted to be alone with me; it was not the fact that no people were trying to reach out or help me.”* SJ

##### 2.1 Barrier to utilising healthcare services: Stigma surrounding depression

There was a general stigma associated with depression[55, 57, 60, 61, 64, 69–71, 73, 76, 77, 83, 94]:

> *“… anything to do with mental health still seems to have that stigma attached where people don’t want to talk about it… so even though you might recognise it and point it out to them, they either ignore what you’re saying or completely deny it…”* [69]

There was also a stigma specific to the Black community[28, 54, 58, 60, 61, 63, 65, 66, 69–71, 73, 75, 77, 79, 82, 83, 90, 91, 93, 94]:

> *“Depression is less accepted in the Black community. Because people just don’t have the patience … you know. They say, “You crazy,” and forget ya.”* [71]

> *“There is a huge stigma when talking about depression. Some disregard my experience because poor mental health is a bad thing to talk about, so we know the stigma exists. People do not want to see you in a weaker light.”* SW

> *“I do not have any concerns about the stigma. We need to have open conversations and speak more about depression. We need to be open and transparent. Previous generations did not have that option, but we are in a position now where we have many platforms.”* SJ

Women feared they would be misunderstood and labelled as ‘crazy’[60, 63, 65, 66, 70, 71, 75, 79, 83, 94] or weak[93, 94]. Therefore, women only disclosed their diagnosis to family[58, 61, 69, 73, 88, 89], friends[56, 58, 61, 65, 66, 69, 83, 86, 89], and romantic partners[89] they trusted for support[28, 54, 56, 58, 60, 61, 65, 66, 69, 73, 77, 79, 83, 86–89, 92]. Some women chose to keep their diagnosis inside the family, which was associated with a cultural belief that others should not know about “family business”[58, 61, 66, 70, 71, 73, 79, 82, 90, 91]. However, some friends and families were not as understanding. Hence, women kept their diagnoses to themselves as they feared their friends and families would not treat them the same[28, 66, 79, 85].

Women who could not disclose their experiences of depression due to stigma and those who had supportive family and friends also engaged in self-help. Some self-help methods such as gardening, sleeping, exercise, writing, meditation, listening to music, watching television, socialising, cleaning, arts and crafts, and talking to themselves were helpful and temporarily alleviated their symptoms[54, 61, 62, 65, 66, 81–83, 87–89]. Other self-help methods such as substance abuse, overeating, overspending, and self-harm (cutting) were not helpful and damaged women’s health[28, 58, 59, 61–63, 66, 76, 79]:

> *“With the self-help, I think that is why Black wellness is a huge thing. We have needed to create our own spaces because we have not been able to find support through traditional mental health care.”* FA

#### 3. Experience: Navigating depression within a framework of faith

As religion was considered historically and culturally embedded in the black community[69, 90–92], women who experienced depression as a loss of faith felt ashamed[54, 62, 69, 73, 77, 79, 80, 84, 86, 95]:

> *“I think that spiritual piece holds more weight in the Black community and even more for middle-class Black women because we have so much to deal with. And just because of our history of our race… a lot of it is connected to religion and our faith.”* [90]

> *“Sometimes I feel ashamed because I feel that God is blessing me each day. So I ask God, ‘What are you trying to tell me? What am I doing that’s not right? What should I be doing?”*[54]

> *“I felt I needed to be part of the church because of what had been drilled into me from a young age. There were many times when I felt my mental health would not improve if I did not tell my pastor about my life or open the Bible to find a chapter relating to whatever I was going through.”* FA

> *“We did not speak about mental health in my household. When I did talk about it, I was told that I had some form of spiritual problem. My parents treated me for that spiritual problem*.” SJ

##### 3.1 Barrier to utilising healthcare services: Religious coping

To combat depression associated with a loss of faith, women utilised religious coping instead of trying to access formal healthcare services for mental health support or treatment[28, 53, 54, 56, 58, 60, 61, 63–66, 69, 71, 77, 79–84, 86–90, 92, 96]. Religious coping included prayer[28, 53, 58, 62–65, 67, 71, 80, 82, 86, 87, 89, 90, 92], reading the Bible [65, 82, 87, 89, 90, 92], speaking to their faith leader [58, 69, 73, 86, 89, 92], and praise and worship[86, 89]:

> *“Nope. I usually depend on my spiritual walk, you know, scriptures, songs to refresh me, bring me out of that state of mind. I don’t take medication…”*[65].

> *“Religious coping is another significant obstacle if you are raised in a religion, and it is not something you subscribe to later in life. It is almost another stigma in your community. However, there have been times when religion was helpful.*” FA

> *“My family would use religion to prevent me from accessing services by saying they would pray for me or give it to God instead of encouraging me to go to a doctor. Those big statements were a barrier for me.”* SW

> *“Initially, I was quite angry with my parents for thinking my depression was a spiritual problem, but in my reflections, I realised that is what they knew. There was some lack of teaching from that aspect.”* SJ

Religion allowed Black women to have faith[56, 60–66, 82–84, 89] that what they were experiencing was short-lived, and they would eventually recover as “God never gives you more trouble than you can bear”[83]. Religious coping was primarily associated with managing symptoms of depression stemming from the oppressions associated with race and gender identities[88].

##### 3.2 Facilitator to utilising healthcare services: Endorsement and support from faith leaders

To utilise formal healthcare services, some women sought approval from their faith leader to feel they were not betraying their faith[73]. Women did not stop believing in God when they eventually utilised services, as they believed that God worked through medicine. Hence, their faith played a significant role in their recovery[74, 84]:

> “*Once I spoke to my pastor, she helped me see that it was OK to get help from a therapist.**”*** [73]

> *“In some churches, going to a faith leader might result in not receiving help and being shunned even more, depending on the level of experience of mental health that the faith leader has. People who have gone against or stepped away from the religion have been exiled from the community, which is crazy. Hence, I think it is something that makes it a lot more difficult for people to navigate when it comes to mental health issues.”* FA

> *“I have family members who are faith leaders. They are so influenced by faith that they disregard medical interventions as helpful. That is the story that resonated with me more.”* SW

> *“Sometimes, there is a hierarchy within the church, so before seeing the leader, you might be asked to read scriptures and complete a 21-day fast.”* SJ

#### 4. Experience: Strong Black woman schema masking depression symptoms

Some women experienced depression as physical symptoms: general pain[54, 63, 86, 88], a tight chest[66, 73], hair loss[66], colds[66], body aches[66, 75, 83], headaches[61, 64, 66], and vaginal pain[61]. Other women experienced typical depression symptoms listed in diagnostics manuals: anger[28, 59, 61–63, 65, 76, 82, 83, 86, 95], sadness[59, 62–64, 66, 75, 79, 86, 92, 95], crying[55, 64, 76, 82, 86, 88], guilt[59, 76, 86, 87], insomnia[55, 64, 73, 76, 86], loss in appetite[76], worthlessness[54, 65, 76, 86, 92, 95], hopelessness[54, 56, 59, 61, 63, 66, 76, 88, 95], ruminating[54, 66, 83, 87], hypersomnia[83], low self-esteem[87], excessive worrying[82], suicidal ideation[59, 86, 88], loss of energy[59, 63, 75], and anhedonia[59, 86]. Although women experienced symptoms, they did not realise they were experiencing depression[63, 89]:

> *“[I] felt stressed but did not realise I was depressed.”* [63]

It was only when their functionality was compromised[54, 61–65, 73, 76, 82, 83, 86, 87, 92, 95] and they could not meet the Strong Black Woman expectation of being able to overcome adversity that it became apparent that they were experiencing depression[54, 61–65, 68, 73, 76, 82, 83, 86, 87, 92, 95]. A loss in functionality was associated with struggling to cope with everyday stressors[28, 56, 57, 61–64, 73, 75, 80, 83, 86, 91, 96]. Sources of stress included looking after family[61, 64, 73, 75, 80, 82, 86, 91], living in socially challenging neighbourhoods[62, 82], financial problems[57, 80], migration[75], and death of family members[63, 64, 73]. Due to the decrease in independence, ability to cope with taking care of others, and hiding their emotions[28, 55], women felt weak, guilty, ashamed, and hopeless as they could not see an escape[28, 93, 94]:

> *“I could almost feel myself shrinking. So that was really, hard, that was really hard, and I still remember it today. There was always little comments like that, letting me know that I wasn’t living up to these women’s definitions of being strong. Of being, you know, being able to take care of things.”* [28]

> *“I think the strong Black woman schema affects you because of the respectability of being a Black woman. We are taught to work twice as hard in school when we are younger and to survive or thrive in society as adults.”* SW

> *“Some women thrive as strong Black women and would not want to be weak or vulnerable. Therefore, it is less about being forced to be the strong Black woman and more about women being forced into a box that is not themselves. If we leave those spaces, everyone will fall apart, so we stay to prevent the guilt of leaving those spaces where we are seen as the glue holding everyone together.”* FA

> *“I feel I am not where I should be because of not being a strong black woman, as I have got a diagnosis. There is a label that I am not a good mother and grandma because we have never had mental health issues in the family until now.”* SJ

##### 4.1 Barrier to utilising healthcare services: Pressure to conform to being a Strong Black Woman

Women denied that they were experiencing depression[28, 57, 64, 66, 71, 73, 76, 78, 82, 83] by internalising their feelings[28, 53, 82, 87] and masking how they felt[58, 59, 61, 63, 66, 69, 71]. Women tried to continue as normal[28, 58, 61, 63–66, 73, 83, 90] as they believed circumstances could be worse[59, 83]:

> *“Somebody’s worser off than we are, so we just got to deal. So that’s where the mask came in. ‘I am a strong Black woman’, so I got to be strong and inside you’re breaking down.”* [59]

The denial of difficult circumstances was associated with attempting to conform to being a Strong Black Woman[28, 53, 54, 57–63, 65, 66, 68–71, 73, 76, 77, 79, 80, 82, 83, 87, 89, 91, 92]. Some women also tried to continue with their caretaking responsibilities[28, 53, 55, 56, 58, 61–64, 71, 73, 82, 88, 91] as they felt obligated[28, 53, 55, 58, 61–63, 68, 73, 91] since their families depended on them. Caring for others became a way to cope with their symptoms as it kept their minds occupied[63, 64, 88]. As the needs of others occupied them, they could disregard their symptoms[71, 82]. When women finally acknowledged that they were unwell, it was hard for them to ask for help[58, 89], so they continued trying to conform to SBWS until they were in crisis:

> *“The strong Black woman schema as a barrier to accessing health interventions is critical. It could link to all the different themes.”* SW

> *“Black women’s experiences have been shaped when we have had negative experiences when we have been vulnerable. We developed a schema that tells us the world is unsafe. However, it is rather the people we have been vulnerable to who were unsafe.”* FA

> *“There have been a lot of comparisons among Black women because of the schema; we look at the strong Black woman and what her counterparts are doing. If their counterparts are doing less than the strong Black woman, then they look like a failure.”* SJ

##### 4.2 Facilitator to utilising healthcare services: Hitting a point of crisis leading to understanding depression

Reaching a crisis point was associated with women realising they needed to care for themselves[58, 59, 62, 63, 66, 69, 73, 79, 89]:

> *“I had to set guidelines for me and for people that counted on me. If you do not make you a priority, nobody is going to. After breaking down, my whole mantra now is … it is all about me.”* [63]

After experiencing a mental health crisis, women realised that depression was not something they could control[62, 70, 73, 76], minor sadness[58, 66, 82, 91], a change in mood[57, 87], associated with social class[61, 79, 82, 91], only influenced by family history[58, 73, 76], or a representation of their Blackness or womanhood[28, 57, 59, 63, 65, 66, 69–71, 73, 76, 77, 79, 82, 83, 88, 90, 91]:

> *“I think with Caucasians, it’s easier for them to admit that they’re depressed, but for African-Americans, there’s more of a sense of a struggle within the community, and to be depressed for what seems like no reason seems almost shameful.”* [70]

While some of the assumptions women held about depression were congruent with those held by clinicians, women highlighted that the lack of information about depression was a barrier to early intervention[54, 57, 58, 60–64, 69, 71, 74, 76]:

> *“Black people don’t want treatment. I think because they not educated about it, you know how important it is … I don’t think they’re informed. People don’t tell them.”* [71]

> *“They [clinicians] do not understand that depression to us is that we are standing on the edge of a cliff. Some of us do not see the lead-up of the symptoms because we have had to navigate so much trauma or harm at a different age or throughout our entire lives, so we have become used to navigating life with depression. So, they think we do not understand what depression is; however, they do not understand how normalised depression is in our lives.”* SW

> *“It is difficult to know what depression is and what is not depression, especially when you have had traumatic experiences. To understand what the recovery, healing and grieving process is supposed to look like. Some say, “It will be better if you talk about it”. However, they do not realise that many people cannot because the people around them are unsafe to talk to.”* FA

## Discussion

### Summary of findings

In this QES, we systematically identified and connected Black women’s experiences of depression with help-seeking behaviours, which were either barriers or facilitators to utilising healthcare services through the construction of a logic model (Figure 3). The model suggests that adverse life experiences, which include racial discrimination experienced by Black women, can promote depression. However, because of the SBWS, women minimise and fail to recognise the significance of their symptoms. Mistrust of HCPs is a barrier to utilising healthcare services, influenced by the relationship between women and HCPs. Stigma is associated with social isolation, exacerbating experiences of depression. Religious coping provides temporary relief but is insufficient for long-term recovery. Seeking support from faith leaders becomes crucial, but without endorsement, women remain confined in the cycle of striving to conform to the SBWS. A crisis, such as a mental health breakdown, can serve as a turning point, prompting recognition of depression and seeking treatment. While the SBWS has traditionally been viewed as comprising three components (i.e., strength, self-reliance, and caretaking and selflessness)[29–31], our research indicates that concerning depression, all the experiences we identified align with elements of the SBWS. Conforming to the SBWS prevents women from engaging in social interactions when depressed, as they may appear weak, and reaching out to others who are part of a faith group, as they do not want to appear immoral. Before conducting our study, we found no systematic reviews and evidence syntheses on Black women’s experiences of depression and barriers and facilitators to healthcare service utilisation.

**Figure 3:**
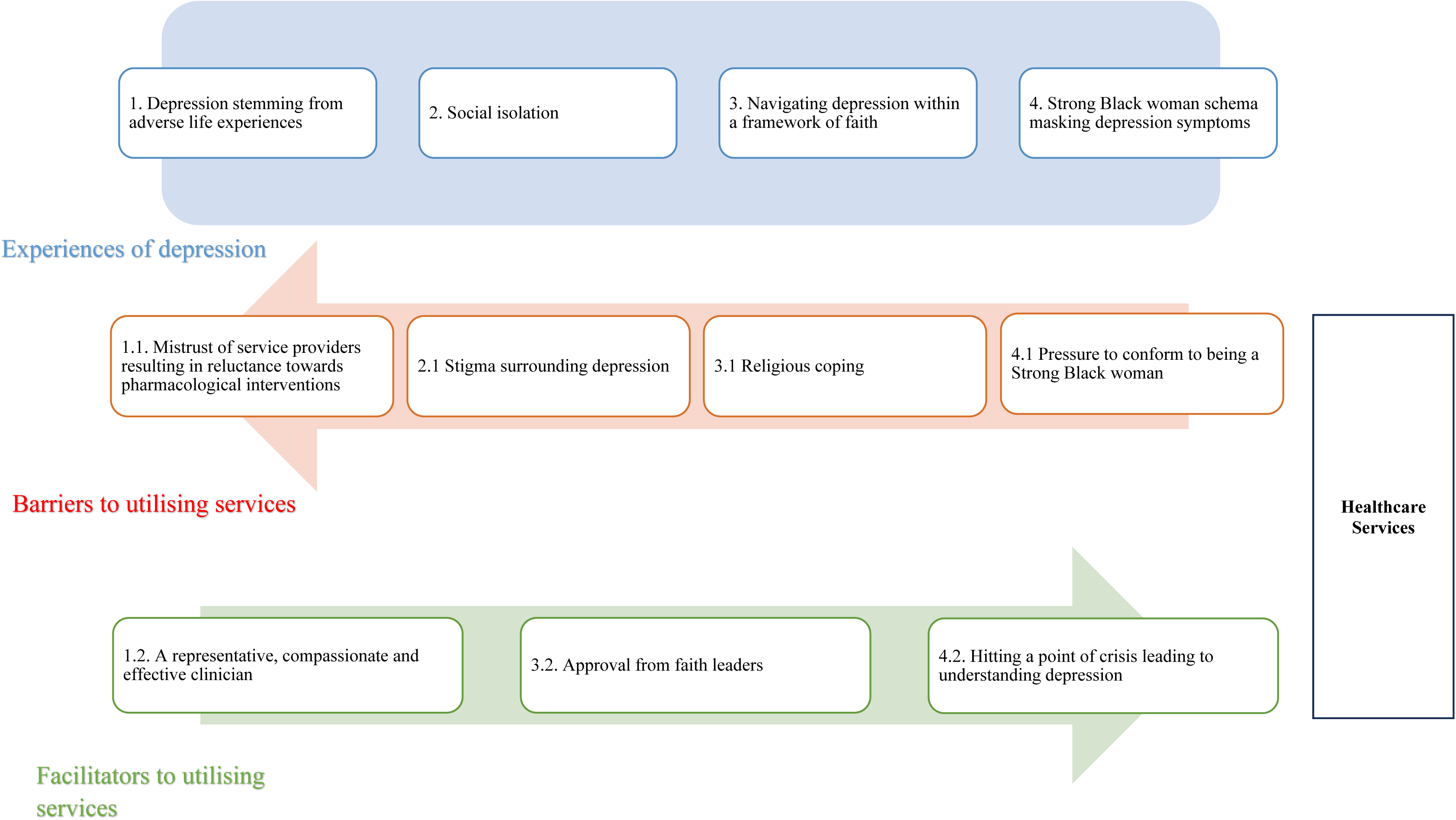
Logic model.

### Main findings with clinical implications

Experiencing depression because of discrimination in the workplace is consistent with findings that token individuals are prone to be mistreated and undervalued[98], and hyper-visibility and invisibility[99].

Hypervisibility exposes individuals to heightened scrutiny and the reinforcement of negative stereotypes, while invisibility denies them recognition and validation for their contributions. Being undervalued and mistreated encourages token individuals to work harder to prove themselves[99]. However, the lack of reciprocity and recognition from non-token individuals often leads to psychological exhaustion and depression[100, 101]. This suggests that psychotherapists should consider the impact of race-related stressors and trauma in and out of the workplace and follow provided guidance (e.g., acknowledge racism as violence and offer empowerment strategies) when treating Black women[102].

Studies that have compared differences between Black and White patients found that greater mistrust among Black individuals[103–105] was rooted in prior personal and vicarious racial discrimination[104]. Furthermore, perceived racism was suggested to have a significant direct effect on satisfaction and cultural mistrust[106]. Hence, mistrust might serve as a protective response to anticipated racial discrimination[107] among Black women. Our findings are aligned with other studies that suggest mistrust is associated with lower healthcare utilisation[108]. Consequently, building trust between HCPs and patients is essential to effective care. For psychotherapists, the therapeutic relationship is formed in the first few sessions. Therefore, all psychotherapists should be well-versed in discussing issues of race to create a trusting therapeutic alliance with Black women[102]. Some training programmes have started incorporating antiracist competencies in training programmes for mental health professionals to teach them how to discuss issues of race[109]. Incorporating antiracist competencies in the curriculums should be mandatory to promote mental health equity and provide equitable access to care for all individuals, regardless of their racial or ethnic background.

Regarding symptoms and the SBWS, our findings suggest that Black women often experience various conventional and unconventional symptoms of depression resonating with QESs findings across the world[42, 110]. However, a novel finding is that to conform to the SBWS, Black women do not recognise that they are depressed. This finding aligns with work by Liao and colleagues[111], which suggests that Black women who endorsed the SBWS are more susceptible to depression, with maladaptive perfectionism acting as a mediator. When Black women felt the pressure to conform to societal expectations of unwavering strength and resilience but fell short of these standards, their likelihood of experiencing depression increased. Liao and colleagues[111] found that self-compassion served as a mediator between the endorsement of the SBWS and depression, meaning women who endorsed the SBWS were more inclined to postpone self-care. Clinicians should be taught to recognise the diverse ways in which depression can manifest, extending beyond the descriptions in diagnostic manuals and conventional treatment. Recent studies suggest that culturally adapted mental health interventions are more effective than conventional approaches[112, 113]. Studies are also advocating for personalised management at the individual level to reflect the heterogeneous and subjective manifestations of depression[114].

The positive psychological change in conceptions concerning Black women’s identity, well-being and depression after experiencing a mental health crisis can be explained by posttraumatic growth (PTG) theory[115]. According to PTG theory, individuals who experience a crisis eventually engage in deliberate and reflective rumination to understand their experiences[115]. The rumination deconstructs or restructures the individuals’ schemas about themselves and life, ultimately leading to personal growth, such as a greater appreciation of life, enhanced intra and interpersonal relations, and a new purpose for their life[115, 116]. Although PTG is typically a natural process occurring without intervention, psychotherapists can facilitate it by integrating PTG principles into their current evidence-based practices[117]. These principles include providing psychoeducation about distress being experienced, helping the patient develop effective emotional regulation techniques to manage intrusive rumination, and encouraging the patient to explore the implications the event has had on their core beliefs, leading to acceptance and growth. Since PTG-based interventions are embedded in cognitive–behavioural, narrative, existential, and interpersonal approaches, they should complement rather than replace existing evidence-based therapies[117].

### Strengths and limitations

This review has several strengths. We developed a logic model linking specific experiences of depression to barriers and facilitators to utilising services. This logic model can be used in future research to develop and test hypotheses and interventions[118] to increase service utilisation and identify and manage depression among this population. Additionally, we consulted experts by lived experience during the research question development, analysis and interpretation, asked experts to provide reflections and recommendations for policy and practice (Table 2), and invited them to co-author the review. This approach facilitates dialogue between experts with lived experience in clinical psychiatric and psychological practice and research, ensuring that findings and recommendations are rooted in the needs of individuals who have lived through these experiences[42]. Finally, our inclusion criteria encompassed clinical and community settings, recognising the low service utilisation[9, 19–21]. Consequently, our findings encompass diverse samples, reflecting a range of experiences and providing a more comprehensive understanding of the barriers faced by the population.

**Table 2:**
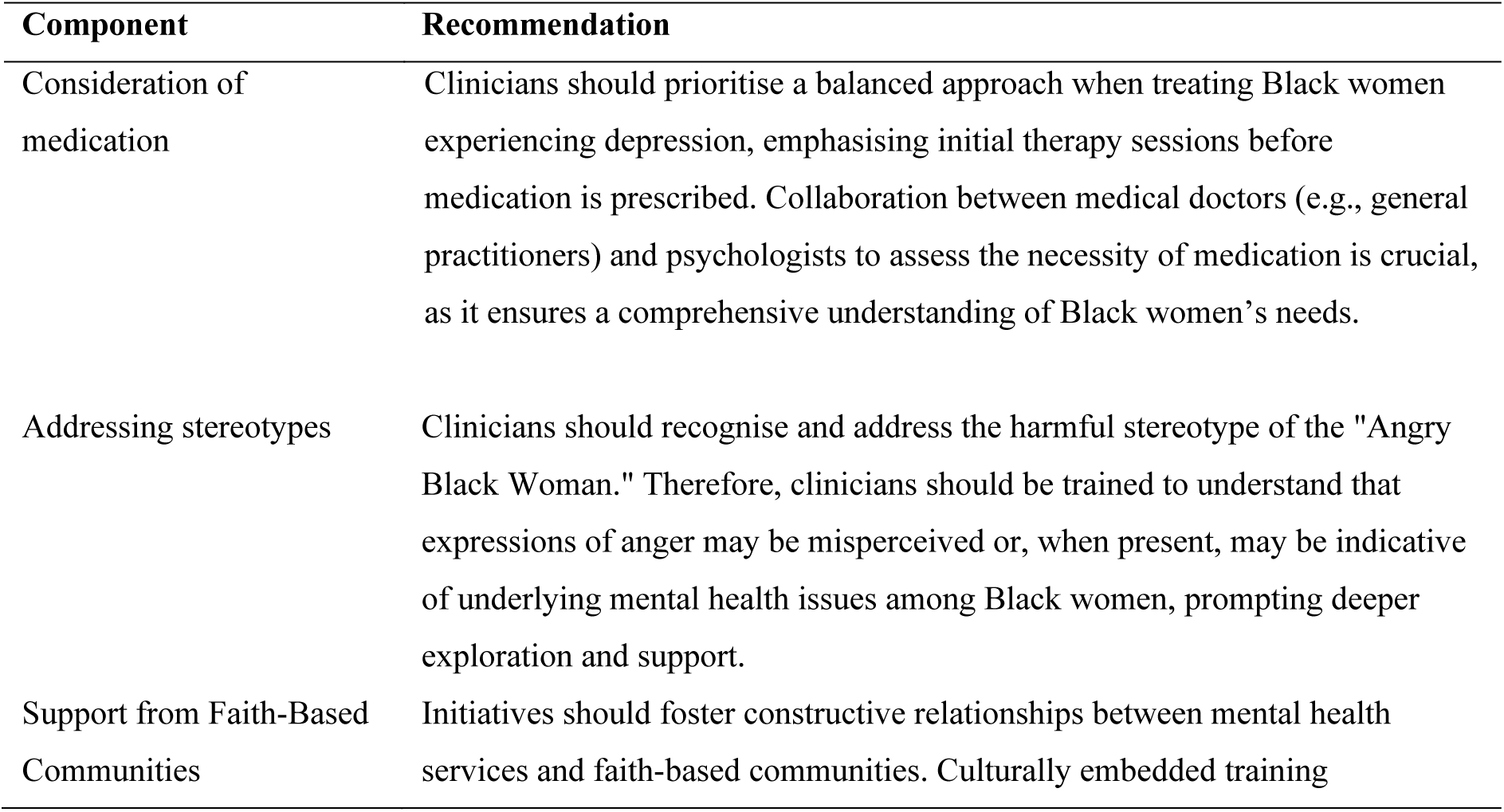
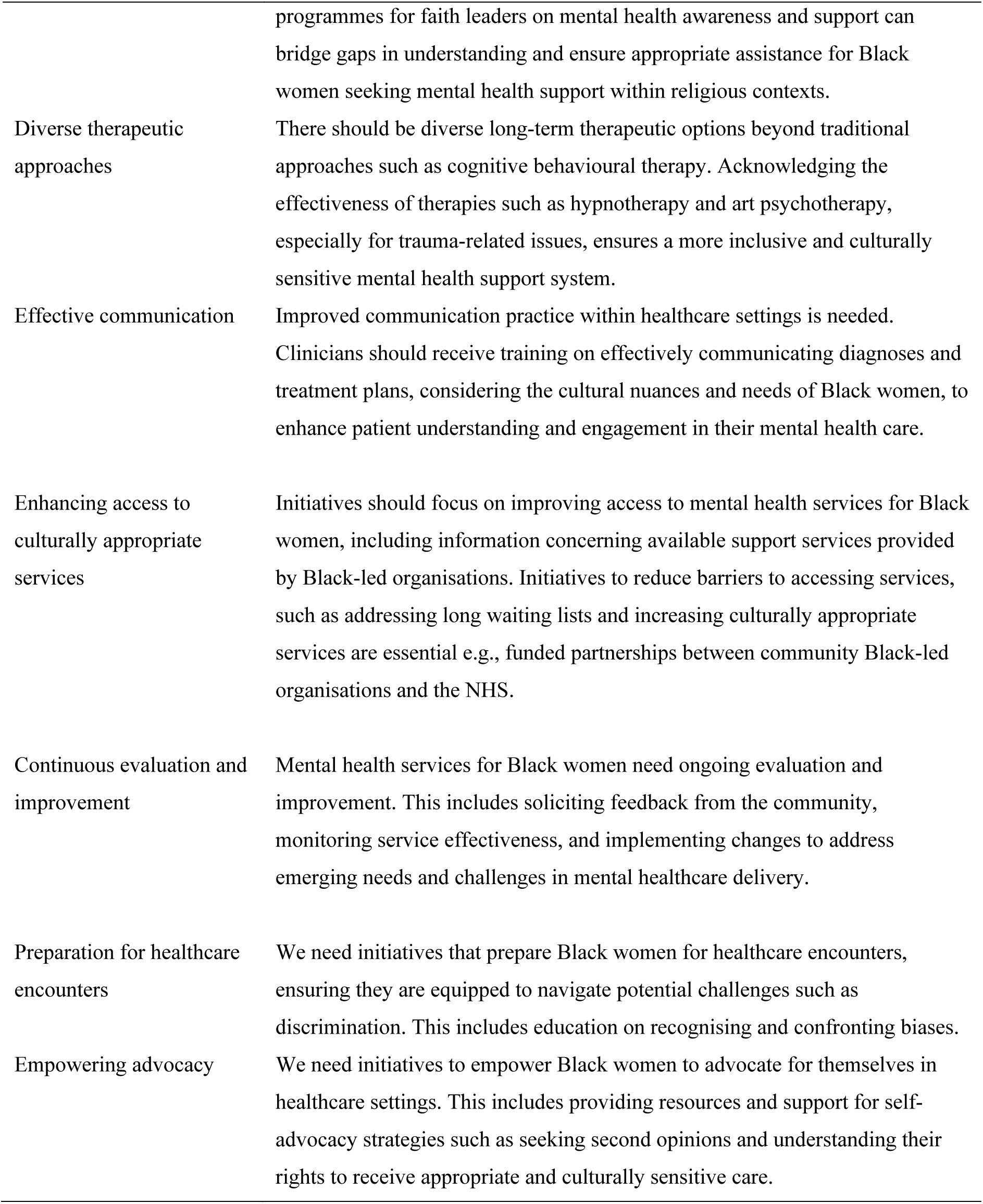
Recommendations for policy and practice from experts by lived experiences.

| **Component** | **Recommendation** |
| --- | --- |
| Consideration of medication | Clinicians should prioritise a balanced approach when treating Black women experiencing depression, emphasising initial therapy sessions before medication is prescribed. Collaboration between medical doctors (e.g., general practitioners) and psychologists to assess the necessity of medication is crucial, as it ensures a comprehensive understanding of Black women's needs. |
| Addressing stereotypes | Clinicians should recognise and address the harmful stereotype of the "Angry Black Woman." Therefore, clinicians should be trained to understand that expressions of anger may be misperceived or, when present, may be indicative of underlying mental health issues among Black women, prompting deeper exploration and support. |
| Support from Faith-Based Communities | Initiatives should foster constructive relationships between mental health services and faith-based communities. Culturally embedded training |
|  | programmes for faith leaders on mental health awareness and support can bridge gaps in understanding and ensure appropriate assistance for Black women seeking mental health support within religious contexts. |
| Diverse therapeutic approaches | There should be diverse long-term therapeutic options beyond traditional approaches such as cognitive behavioural therapy. Acknowledging the effectiveness of therapies such as hypnotherapy and art psychotherapy, especially for trauma-related issues, ensures a more inclusive and culturally sensitive mental health support system. |
| Effective communication | Improved communication practice within healthcare settings is needed. Clinicians should receive training on effectively communicating diagnoses and treatment plans, considering the cultural nuances and needs of Black women, to enhance patient understanding and engagement in their mental health care. |
| Enhancing access to culturally appropriate services | Initiatives should focus on improving access to mental health services for Black women, including information concerning available support services provided by Black-led organisations. Initiatives to reduce barriers to accessing services, such as addressing long waiting lists and increasing culturally appropriate services are essential e.g., funded partnerships between community Black-led organisations and the NHS. |
| Continuous evaluation and improvement | Mental health services for Black women need ongoing evaluation and improvement. This includes soliciting feedback from the community, monitoring service effectiveness, and implementing changes to address emerging needs and challenges in mental healthcare delivery. |
| Preparation for healthcare encounters | We need initiatives that prepare Black women for healthcare encounters, ensuring they are equipped to navigate potential challenges such as discrimination. This includes education on recognising and confronting biases. |
| Empowering advocacy | We need initiatives to empower Black women to advocate for themselves in healthcare settings. This includes providing resources and support for self-advocacy strategies such as seeking second opinions and understanding their rights to receive appropriate and culturally sensitive care. |

We acknowledge some limitations of this QES. Most studies were conducted in the USA; therefore, experiences, barriers, and facilitators may reflect health services in the USA, limiting our understanding due to differences in healthcare systems. However, this might not be the case as a recent QES in the UK exploring barriers to mental health support among racially minoritised individuals also reported stigma, understanding distress and coping, and competence of the HCPs as barriers[119]. Another challenge lies in the averaging approach to synthesising experiences. Although this QES was comprehensive and provided heterogeneous subjective experiences, some findings may not resonate with all Black women. Finally, our review’s exclusive inclusion of grey literature in the form of PhD theses published in databases and papers published in English may have resulted in the inadvertent omission of relevant studies and reports from health services, charities, or third-sector organisations. These unexplored sources could have offered valuable insights into the subject.

### Future research

In future research, better reporting of participants’ demographic information is needed in qualitative studies. Thirteen studies were excluded from this review as it was unclear which excerpts belonged to Black women. These studies often used homogenising terms such as ethnic minority, underserved or Black Asian and Minority Ethnic (BAME). Understanding the unique experiences of depression and help-seeking behaviours within different racial groups hinges on the reporting of complete demographic data. Black women’s history of maltreatment has played a significant role in Black women not being able to trust HCPs. Hence, future research should focus on developing and evaluating culturally adapted interventions for Black women. These interventions should incorporate cultural humility and competence from HCPs to foster trust and long-term engagement in healthcare services[120, 121]. Although cultural competence and humility have been considered opposing approaches, others focusing on experiences of depression among Black women contend that they can complement each other[120]. By incorporating both into practice, HCPs might be able to tackle both micro-level issues by centring the patient’s experiences through humility and macro-level issues through cultural competency. Importantly, to our knowledge, there is no quantitative research in the UK to support that Black British women endorse the SBWS. However, from our findings, qualitative research indicates this might be the case[61, 66, 96]. Hence, future research in the UK might benefit from investigating whether Black British women endorse the SBWS and whether its endorsement predicts adverse outcomes.

## Conclusion

By synthesising evidence from 45 studies in collaboration with experts by lived experiences, this review provides richer and more nuanced findings concerning the lived experiences of depression among Black women and related barriers and facilitators to utilising healthcare services compared with previous research. Overall, recognition and acknowledgement of depression and help-seeking may be hampered by intersectional schemas (i.e., SBWS), which are deeply connected to Black women’s identity. Our findings can guide future research to develop and test culturally tailored interventions for identifying and treating depression, ultimately increasing the use of healthcare services by Black women.

## Data Availability

All data produced in the present work are contained in the manuscript.

## Contributors

AJ was the lead researcher responsible for the study design, screenings, quality appraisals, data extraction, results synthesis, and manuscript writing. JJ and KB supervised and provided input on the study design, screenings, quality appraisals, data extraction, results synthesis, and manuscript writing. FS screened titles and abstracts, full text, quality appraised and supported with interpretations of data. KY supported the analysis and synthesis of the results and PPIE activities. At the start of the project, JO was a supervisor and in the PPIE advisory group, supporting the design, analysis, synthesis, and interpretation of the data. FA, SW, and SJ supported the formulation of the research question, data analysis, synthesis, and interpretation of the data. All authors provided comments on drafts of the manuscript.

## Data sharing

Data will be made available upon request.

## Declaration of interest

We declare no competing interests.

## Acknowledgements

We thank Isabelle Butcher for reviewing the manuscript drafts and offering insight.

## Funding

The Economic Social Research Council funded AJ via the London Interdisciplinary Social Science Doctoral Training Partnership. Grant award number: ES/P000703/1 – PR: 2462475. The PPIE activities were funded by the Centre of Public Engagement at Queen Mary University of London and the National Institute for Health Research Applied Research Collaborative (ARC) North Thames through grants held by AJ. The views expressed in this publication are those of the author(s) and not necessarily those of the National Institute for Health Research or the Department of Health and Social Care. KB is partly supported by Oxford Health NIHR BRC and Thames Valley & Oxford NIHR Applied Research Collaborative.

## Notes

### Competing Interest Statement

The authors have declared no competing interest.

### Author Declarations

This is a systematic review and qualitative evidence synthesis, so we used data from published papers. The papers are cited and referenced. Therefore, data can be accessed from them.

