## Supplemental Materials for "Black Women’s Lived Experiences of Depression and Related Barriers and Facilitators to Utilising Healthcare Services: A Systematic Review and Qualitative Evidence Synthesis Co-produced with Experts by Lived Experiences"

### Supplementary Appendices

#### Table of Contents of Supplementary Appendix

**Appendix A:** Updated Preferred Reporting Items or Systematic Reviews and Meta-Analyses Statement Guidelines

| Section and Topic | Item # | Checklist item | Location where item is reported |
|---|---|---|---|
| **TITLE** | | | |
| Title | 1 | Identify the report as a systematic review. | Manuscript (MS) Title page |
| **ABSTRACT** | | | |
| Abstract | 2 | See the PRISMA 2020 for Abstracts checklist. | MS Abstract |
| **INTRODUCTION** | | | |
| Rationale | 3 | Describe the rationale for the review in the context of existing knowledge. | MS introduction |
| Objectives | 4 | Provide an explicit statement of the objective(s) or question(s) the review addresses. | MS objectives |
| **METHODS** | | | |
| Eligibility criteria | 5 | Specify the inclusion and exclusion criteria for the review and how studies were grouped for the syntheses. | MS criteria for study selection |
| Information sources | 6 | Specify all databases, registers, websites, organisations, reference lists and other sources searched or consulted to identify studies. Specify the date when each source was last searched or consulted. | MS search methods and appendix C |
| Search strategy | 7 | Present the full search strategies for all databases, registers and websites, including any filters and limits used. | Appendix C |
| Selection process | 8 | Specify the methods used to decide whether a study met the inclusion criteria of the review, including how many reviewers screened each record and each report retrieved, whether they worked independently, and if applicable, details of automation tools used in the process. | MS study selection |
| Data collection process | 9 | Specify the methods used to collect data from reports, including how many reviewers collected data from each report, whether they worked independently, any processes for obtaining or confirming data from study investigators, and if applicable, details of automation tools used in the process. | MS data extraction |
| Data items | 10a | List and define all outcomes for which data were sought. | N/A |

| | | | |
|---|---|---|---|
| | | Specify whether all results that were compatible with each outcome domain in each study were sought (e.g. for all measures, time points, analyses), and if not, the methods used to decide which results to collect. | |
| | 10b | List and define all other variables for which data were sought (e.g. participant and intervention characteristics, funding sources). Describe any assumptions made about any missing or unclear information. | MS data extraction |
| Study risk of bias assessment | 11 | Specify the methods used to assess risk of bias in the included studies, including details of the tool(s) used, how many reviewers assessed each study and whether they worked independently, and if applicable, details of automation tools used in the process. | N/A |
| Effect measures | 12 | Specify for each outcome the effect measure(s) (e.g. risk ratio, mean difference) used in the synthesis or presentation of results. | N/A |
| Synthesis methods | 13a | Describe the processes used to decide which studies were eligible for each synthesis (e.g. tabulating the study intervention characteristics and comparing against the planned groups for each synthesis (item #5)). | MS data management, analysis and synthesis |
| | 13b | Describe any methods required to prepare the data for presentation or synthesis, such as handling of missing summary statistics, or data conversions. | N/A |
| | 13c | Describe any methods used to tabulate or visually display results of individual studies and syntheses. | N/A |
| | 13d | Describe any methods used to synthesize results and provide a rationale for the choice(s). If meta-analysis was performed, describe the model(s), method(s) to identify the presence and extent of statistical heterogeneity, and software package(s) used. | N/A |
| | 13e | Describe any methods used to explore possible causes of heterogeneity among study results (e.g. subgroup analysis, meta-regression). | N/A |
| | 13f | Describe any sensitivity analyses conducted to assess robustness of the synthesized results. | N/A |
| Reporting bias | 14 | Describe any methods used to assess risk of bias due to missing results in a synthesis (arising from reporting biases). | N/A |

| | | | |
|---|---|---|---|
| assessment | | | |
| Certainty assessment | 15 | Describe any methods used to assess certainty (or confidence) in the body of evidence for an outcome. | MS assessing confidence in the findings |
| **RESULTS** | | | |
| Study selection | 16a | Describe the results of the search and selection process, from the number of records identified in the search to the number of studies included in the review, ideally using a flow diagram. | MS figure 2 |
| | 16b | Cite studies that might appear to meet the inclusion criteria, but which were excluded, and explain why they were excluded. | N/A |
| Study characteristics | 17 | Cite each included study and present its characteristics. | Appendix D |
| Risk of bias in studies | 18 | Present assessments of risk of bias for each included study. | N/A |
| Results of individual studies | 19 | For all outcomes, present, for each study: (a) summary statistics for each group (where appropriate) and (b) an effect estimate and its precision (e.g. confidence/credible interval), ideally using structured tables or plots. | N/A |
| Results of syntheses | 20a | For each synthesis, briefly summarise the characteristics and risk of bias among contributing studies. | N/A |
| | 20b | Present results of all statistical syntheses conducted. If meta-analysis was done, present for each the summary estimate and its precision (e.g. confidence/credible interval) and measures of statistical heterogeneity. If comparing groups, describe the direction of the effect. | N/A |
| | 20c | Present results of all investigations of possible causes of heterogeneity among study results. | N/A |
| | 20d | Present results of all sensitivity analyses conducted to assess the robustness of the synthesized results. | N/A |
| Reporting biases | 21 | Present assessments of risk of bias due to missing results (arising from reporting biases) for each synthesis assessed. | N/A |
| Certainty of evidence | 22 | Present assessments of certainty (or confidence) in the body of evidence for each outcome assessed. | MS table 1 and appendix F |
| **DISCUSSION** | | | |
| Discussion | 23a | Provide a general interpretation of the results in the context of | MS discussion |

| | | | |
|---|---|---|---|
| | | other evidence. | |
| | 23b | Discuss any limitations of the evidence included in the review. | MS strength and limitations, Methodological limitations of the studies and appendix G |
| | 23c | Discuss any limitations of the review processes used. | MS strength and limitations |
| | 23d | Discuss implications of the results for practice, policy, and future research. | MS table 2 and Discussion |
| **OTHER INFORMATION** | | | |
| Registration and protocol | 24a | Provide registration information for the review, including register name and registration number, or state that the review was not registered. | MS methods |
| | 24b | Indicate where the review protocol can be accessed, or state that a protocol was not prepared. | MS methods |
| | 24c | Describe and explain any amendments to information provided at registration or in the protocol. | In the protocol on PROSPERO |
| Support | 25 | Describe sources of financial or non-financial support for the review, and the role of the funders or sponsors in the review. | MS acknowledgements and funding |
| Competing interests | 26 | Declare any competing interests of review authors. | N/A |
| Availability of data, code and other materials | 27 | Report which of the following are publicly available and where they can be found: template data collection forms; data extracted from included studies; data used for all analyses; analytic code; any other materials used in the review. | MS data sharing |

#### Appendix B: Enhancing transparency in reporting the synthesis of qualitative research (ENTREQ) checklist

| No | Item | Guide and description | Page # |
|---|---|---|---|
| **1** | Aim | State the research question the synthesis addresses. | 2 |
| **2** | Synthesis methodology | Identify the synthesis methodology or theoretical framework which underpins the synthesis and describe the rationale for choice of methodology *(e.g. meta-ethnography, thematic synthesis, critical interpretive synthesis, grounded theory synthesis, realist synthesis, meta-aggregation, meta-study, framework synthesis).* | 4 |
| **3** | Approach to searching | Indicate whether the search was pre-planned (*comprehensive search strategies to seek all available studies)* or iterative (*to seek all available concepts until they theoretical saturation is achieved).* | 3 |
| **4** | Inclusion criteria | Specify the inclusion/exclusion criteria *(e.g. in terms of population, language, year limits, type of publication, study type).* | 3 |
| **5** | Data sources | Describe the information sources used (e.g. *electronic databases (MEDLINE, EMBASE, CINAHL, psycINFO, Econlit), grey literature databases (digital thesis, policy reports), relevant organisational websites, experts, information specialists, generic web searches (Google Scholar) hand searching, reference lists)* and when the searches conducted; provide the rationale for using the data sources. | 3 |
| **6** | Electronic Search strategy | Describe the literature search *(e.g. provide electronic search strategies with population terms, clinical or health topic terms, experiential or social phenomena related terms, filters for qualitative research, and search limits).* | 3 and appendix C |
| **7** | Study screening methods | Describe the process of study screening and sifting *(e.g. title, abstract and full text review, number of independent reviewers who screened studies).* | 3-4 |
| **8** | Study characteristics | Present the characteristics of the included studies *(e.g. year of publication, country, population, number of participants, data collection, methodology, analysis, research questions).* | 5-6 and appendix D |
| **9** | Study selection results | Identify the number of studies screened and provide reasons for study exclusion *(e,g, for comprehensive searching, provide numbers of studies screened and reasons for exclusion indicated in a figure/flowchart; for iterative searching describe reasons for study exclusion and inclusion based on modifications t the research question and/or contribution to theory development).* | 5 and Figure 2 |
| **10** | Rationale for appraisal | Describe the rationale and approach used to appraise the included studies or selected findings *(e.g. assessment of conduct (validity and robustness), assessment of reporting (transparency), assessment of content and utility of the findings).* | 4 |
| **11** | Appraisal items | State the tools, frameworks and criteria used to appraise the studies or selected findings *(e.g. Existing tools: CASP, QARI, COREQ, Mays and Pope* [25]*; reviewer developed tools; describe the domains assessed: research team, study design, data analysis and interpretations, reporting).* | 4 |

| | | | |
|---|---|---|---|
| **12** | Appraisal process | Indicate whether the appraisal was conducted independently by more than one reviewer and if consensus was required. | 4 |
| **13** | Appraisal results | Present results of the quality assessment and indicate which articles, if any, were weighted/excluded based on the assessment and give the rationale. | 6-8, table 1 and appendices E and F |
| **14** | Data extraction | Indicate which sections of the primary studies were analysed and how were the data extracted from the primary studies? *(e.g. all text under the headings "results /conclusions" were extracted electronically and entered into a computer software).* | 4 |
| **15** | Software | State the computer software used, if any. | 4 |
| **16** | Number of reviewers | Identify who was involved in coding and analysis. | 4 |
| **17** | Coding | Describe the process for coding of data *(e.g. line by line coding to search for concepts).* | 4 |
| **18** | Study comparison | Describe how were comparisons made within and across studies *(e.g. subsequent studies were coded into pre-existing concepts, and new concepts were created when deemed necessary).* | 4 |
| **19** | Derivation of themes | Explain whether the process of deriving the themes or constructs was inductive or deductive. | 4 |
| **20** | Quotations | Provide quotations from the primary studies to illustrate themes/constructs, and identify whether the quotations were participant quotations of the author's interpretation. | 8-16 |
| **21** | Synthesis output | Present rich, compelling and useful results that go beyond a summary of the primary studies (e.g. *new interpretation, models of evidence, conceptual models, analytical framework, development of a new theory or construct).* | 8-16 and figure 3 |

#### Appendix C: Search Strategies

| Date searched | Journal/Database | Search string | Limitations | Total Hits |
|---|---|---|---|---|
| 2.09.21 and updated to 29.03.24 | ASSIA | ab(Black OR Afro-Caribbean OR "African American" OR "African Caribbean" OR "West Indian" OR "Black African" OR Coloured OR "Women of colour" OR "Strong Black")OR pub(Black OR Afro-Caribbean OR "African American" OR "African Caribbean" OR "West Indian" OR "Black African" OR Coloured OR "Women of colour" OR "Strong Black")OR mainsubject(Black OR Afro-Caribbean OR "African American" OR "African Caribbean" OR "West Indian" OR "Black African" OR Coloured OR "Women of colour" OR "Strong Black")) AND (ab(Women OR Woman OR Female OR Girl/s OR Lady) OR pub(Women OR Woman OR Female OR Girl/s OR Lady) OR mainsubject(Women OR Woman OR Female OR Girl/s OR Lady)) AND (ab(Depression OR "Psychological distress" OR "Low mood" OR "Feeling low" OR Sad OR "Mood disorder" OR "Affective disorder" OR Hopelessness OR Melancholia OR "Depressive symptoms" OR "Emotional Distress" OR Miserable OR "Sisterella complex") OR pub(Depression OR "Psychological distress" OR "Low mood" OR "Feeling low" OR Sad OR "Mood disorder" OR "Affective disorder" OR Hopelessness OR Melancholia OR "Depressive symptoms" OR "Emotional Distress" OR Miserable OR "Sisterella complex") OR mainsubject(Depression OR "Psychological distress" OR "Low mood" OR "Feeling low" OR Sad OR "Mood disorder" OR "Affective disorder" OR Hopelessness OR Melancholia OR "Depressive symptoms" OR "Emotional Distress" OR Miserable OR "Sisterella complex")) | English Language | 1322 |

| | | | | |
|---|---|---|---|---|
| 72.09.21 and updated to 29.03.24 | MEDLINE | (Black or Afro-Caribbean or "African American" or "African Caribbean" or "West Indian" or "Black African" or Coloured or "Women of colour" or "Strong Black").ab. or (Black or Afro-Caribbean or "African American" or "African Caribbean" or "West Indian" or "Black African" or Coloured or "Women of colour" or "Strong Black").ti. or (Black or Afro-Caribbean or "African American" or "African Caribbean" or "West Indian" or "Black African" or Coloured or "Women of colour" or "Strong Black").sh. AND (Women or Woman or Female or Girl s or Lady).ab. OR (Women or Woman or Female or Girl s or Lady).ti. OR (Women or Woman or Female or Girl s or Lady).sh.<br>AND (Depression or "Psychological distress" or "Low mood" or "Feeling low" or Sad or "Mood disorder" or "Affective disorder" or Hopelessness or Melancholia or "Depressive symptoms" or "Emotional Distress" or Miserable or "Sisterella complex").ab. OR (Depression or "Psychological distress" or "Low mood" or "Feeling low" or Sad or "Mood disorder" or "Affective disorder" or Hopelessness or Melancholia or "Depressive symptoms" or "Emotional Distress" or Miserable or "Sisterella complex").ti. OR (Depression or "Psychological distress" or "Low mood" or "Feeling low" or Sad or "Mood disorder" or "Affective disorder" or Hopelessness or Melancholia or "Depressive symptoms" or "Emotional Distress" or Miserable or "Sisterella complex").sh. | English Language and humans | 5989 |
| 2.09.21 and updated to 29.03.24 | APA PsycInfo | (Black or Afro-Caribbean or "African American" or "African Caribbean" or "West Indian" or "Black African" or Coloured or "Women of colour" or "Strong Black").ab. or (Black or Afro-Caribbean or "African American" or "African Caribbean" or "West Indian" or "Black African" or Coloured or "Women of colour" or "Strong Black").ti. or (Black or Afro-Caribbean or | English Language and human | 5006 |

| | | | | |
|---|---|---|---|---|
| | | "African American" or "African Caribbean" or "West Indian" or "Black African" or Coloured or "Women of colour" or "Strong Black").mh. AND (Women or Woman or Female or Girl/s or Lady).ab. or (Women or Woman or Female or Girl/s or Lady).ti. or (Women or Woman or Female or Girl/s or Lady).mh. AND (Depression or "Psychological distress" or "Low mood" or "Feeling low" or Sad or "Mood disorder" or "Affective disorder" or Hopelessness or Melancholia or "Depressive symptoms" or "Emotional Distress" or Miserable or "Sisterella complex").ab. or (Depression or "Psychological distress" or "Low mood" or "Feeling low" or Sad or "Mood disorder" or "Affective disorder" or Hopelessness or Melancholia or "Depressive symptoms" or "Emotional Distress" or Miserable or "Sisterella complex").ti. or (Depression or "Psychological distress" or "Low mood" or "Feeling low" or Sad or "Mood disorder" or "Affective disorder" or Hopelessness or Melancholia or "Depressive symptoms" or "Emotional Distress" or Miserable or "Sisterella complex").mh. | | |
| 2.09.21 and updated to 29.03.24 | Sociological Abstracts | (ab(Depression OR "Psychological distress" OR "Low mood" OR "Feeling low" OR Sad OR "Mood disorder" OR "Affective disorder" OR Hopelessness OR Melancholia OR "Depressive symptoms" OR "Emotional Distress" OR Miserable OR "Sisterella complex") OR mainsubject(Depression OR "Psychological distress" OR "Low mood" OR "Feeling low" OR Sad OR "Mood disorder" OR "Affective disorder" OR Hopelessness OR Melancholia OR "Depressive symptoms" OR "Emotional Distress" OR Miserable OR "Sisterella complex") OR pub(Depression OR "Psychological distress" OR "Low mood" OR "Feeling low" OR Sad OR "Mood disorder" OR "Affective disorder" OR Hopelessness OR Melancholia OR "Depressive | English Language | 1124 |

| | | | | |
|---|---|---|---|---|
| | | symptoms" OR "Emotional Distress" OR Miserable OR "Sisterella complex")) AND (ab(Women OR Woman OR Female OR Girl/s OR Lady) OR mainsubject(Women OR Woman OR Female OR Girl/s OR Lady) OR pub(Women OR Woman OR Female OR Girl/s OR Lady)) AND (ab(Black OR Afro-Caribbean OR "African American" OR "African Caribbean" OR "West Indian" OR "Black African" OR Coloured OR "Women of colour" OR "Strong Black") OR mainsubject(Black OR Afro-Caribbean OR "African American" OR "African Caribbean" OR "West Indian" OR "Black African" OR Coloured OR "Women of colour" OR "Strong Black") OR pub(Black OR Afro-Caribbean OR "African American" OR "African Caribbean" OR "West Indian" OR "Black African" OR Coloured OR "Women of colour" OR "Strong Black")) | | |
| 2.09.21 and updated to 29.03.24 | CINAHL | AB ( Black OR Afro-Caribbean OR "African American" OR "African Caribbean" OR "West Indian" OR "Black African" OR Coloured OR "Women of colour" OR "Strong Black" ) OR TI ( Black OR Afro-Caribbean OR "African American" OR "African Caribbean" OR "West Indian" OR "Black African" OR Coloured OR "Women of colour" OR "Strong Black" ) OR MW ( Black OR Afro-Caribbean OR "African American" OR "African Caribbean" OR "West Indian" OR "Black African" OR Coloured OR "Women of colour" OR "Strong Black" ) AND AB ( Women OR Woman OR Female OR Girl/s OR Lady ) OR TI ( Women OR Woman OR Female OR Girl/s OR Lady ) OR MW ( Women OR Woman OR Female OR Girl/s OR Lady ) AND AB ( Depression OR "Psychological distress" OR "Low mood" OR "Feeling low" OR Sad OR "Mood disorder" OR "Affective disorder" OR Hopelessness OR Melancholia OR "Depressive symptoms" OR "Emotional Distress" OR Miserable OR "Sisterella complex" ) OR TI ( | English Language and human | 4322 |

| | | | | |
|---|---|---|---|---|
| | | Depression OR "Psychological distress" OR "Low mood" OR "Feeling low" OR Sad OR "Mood disorder" OR "Affective disorder" OR Hopelessness OR Melancholia OR "Depressive symptoms" OR "Emotional Distress" OR Miserable OR "Sisterella complex" ) OR MW ( Depression OR "Psychological distress" OR "Low mood" OR "Feeling low" OR Sad OR "Mood disorder" OR "Affective disorder" OR Hopelessness OR Melancholia OR "Depressive symptoms" OR "Emotional Distress" OR Miserable OR "Sisterella complex" ) | | |
| 2.09.21 and updated to 29.03.24 | AMED | (Black or Afro-Caribbean or "African American" or "African Caribbean" or "West Indian" or "Black African" or Coloured or "Women of colour" or "Strong Black").ab. or (Black or Afro-Caribbean or "African American" or "African Caribbean" or "West Indian" or "Black African" or Coloured or "Women of colour" or "Strong Black").ti. or (Black or Afro-Caribbean or "African American" or "African Caribbean" or "West Indian" or "Black African" or Coloured or "Women of colour" or "Strong Black").sh. AND (Women or Woman or Female or Girl/s or Lady).ab. or (Women or Woman or Female or Girl/s or Lady).ti. or (Women or Woman or Female or Girl/s or Lady).sh. AND (Depression or "Psychological distress" or "Low mood" or "Feeling low" or Sad or "Mood disorder" or "Affective disorder" or Hopelessness or Melancholia or "Depressive symptoms" or "Emotional Distress" or Miserable or "Sisterella complex").ab. or (Depression or "Psychological distress" or "Low mood" or "Feeling low" or Sad or "Mood disorder" or "Affective disorder" or Hopelessness or Melancholia or "Depressive symptoms" or "Emotional Distress" or Miserable or "Sisterella complex").ti. or (Depression or "Psychological distress" or "Low mood" or "Feeling low" or Sad or "Mood disorder" or | No limits | 39 |

| | | | | |
|---|---|---|---|---|
| | | "Affective disorder" or Hopelessness or Melancholia or "Depressive symptoms" or "Emotional Distress" or Miserable or "Sisterella complex").sh. | | |
| 2.09.21 and updated to 29.03.24 | EMBASE | (Black or Afro-Caribbean or "African American" or "African Caribbean" or "West Indian" or "Black African" or Coloured or "Women of colour" or "Strong Black").ab. or (Black or Afro-Caribbean or "African American" or "African Caribbean" or "West Indian" or "Black African" or Coloured or "Women of colour" or "Strong Black").ti. or (Black or Afro-Caribbean or "African American" or "African Caribbean" or "West Indian" or "Black African" or Coloured or "Women of colour" or "Strong Black").sh. AND (Women or Woman or Female or Girl/s or Lady).ab. or (Women or Woman or Female or Girl/s or Lady).ti. or (Women or Woman or Female or Girl/s or Lady).sh. AND (Depression or "Psychological distress" or "Low mood" or "Feeling low" or Sad or "Mood disorder" or "Affective disorder" or Hopelessness or Melancholia or "Depressive symptoms" or "Emotional Distress" or Miserable or "Sisterella complex").ab. or (Depression or "Psychological distress" or "Low mood" or "Feeling low" or Sad or "Mood disorder" or "Affective disorder" or Hopelessness or Melancholia or "Depressive symptoms" or "Emotional Distress" or Miserable or "Sisterella complex").ti. or (Depression or "Psychological distress" or "Low mood" or "Feeling low" or Sad or "Mood disorder" or "Affective disorder" or Hopelessness or Melancholia or "Depressive symptoms" or "Emotional Distress" or Miserable or "Sisterella complex").sh. | Human and English Language | 7031 |

#### Appendix D: Table summary of characteristics of included papers

| Author | Research Aim/Questions | Geographical Area | Data Collection Method | Data Analysis Method | Number of Participants | Ethnicity | Age | Were experiences or levels of depression measured as part of the study? | Was current or previous treatment for depression measured as part of the study? |
|---|---|---|---|---|---|---|---|---|---|
| Akinyemi (2018) | Explore the language used by older, church-going African Americans to describe depression and how this language compares to that used by clinicians whose objective is to make clinical diagnoses based on DSM-IV criteria. | US | Focus Groups | Content Analysis | Total 50: Black women 29 | African American | Mean: 67 Range: 51 to 94 | No | Not Stated |
| Alang (2016) | To identify meanings and common ways of expressing depression among a community of African Americans | US | Ethnography | Constant comparative coding analysis | Not stated | African American | Not stated | No | No |

| | | | | | | | | | |
|---|---|---|---|---|---|---|---|---|---|
| | living in a disadvantaged neighbourhood. | | | | | | | | |
| Anderson (2021) | How is psychological distress experienced by African American female mid-level leaders who have undergone intersectional invisibility in large US organisations? | US | Interviews | Interpretative phenomenological analysis | 10 | Black | Range: 30 to 70 | Yes (self-disclosure) | Not Stated |
| Atanmo-Strempek (2014) | What are African American women's experiences related to depression in the African American church? How do African American women view the role of the church in coping with depressive symptoms? | US | Interviews | Content Analysis | 6 | African American | Range: 29 to 60 | Yes (validated questionnaire) The Beck Depression Inventory-II 20 points or higher on the BDI-II to participate | Yes |

| | | | | | | | | | |
|---|---|---|---|---|---|---|---|---|---|
| Bailey (2021) | To explore help-seeking views and strategies utilised in relation to depression among older Black Caribbean people in the UK. | UK | Interviews | Interpretative phenomenologica l analysis | Total 8: Black women 4 | Black Caribbean | Mean: 71.4 Range: 65 to 79 | Not Stated | No |
| Barbee (1994) | The purpose of the present study was to reframe dysphoric mood states experienced by African American women through the lens of anthropology to explore how their cultural orientation influences and transforms what is usually viewed as a pathological state into a vehicle for self-help and renewal. | US | Interviews | Not stated | 15 | African American | Mean: 43 Range: 26 to 57 | No | No |
| Beagan (2012) | To shed light on how racism-related stress, mental health and coping were experienced and perceived by some | Canada Nova Scotia | Interviews | Thematic Analysis (Inductive) | 50 | African-heritage, multi-generation Nova Scotian | Mean: 51 | Yes (validated questionnaire) Centre for Epidemiologic Studies | Not Stated |

| | | | | | | | | | |
|---|---|---|---|---|---|---|---|---|---|
| | African Nova Scotian women, highlighting the role of spirituality as a source of health and well-being. | | | | | | | Depression Scale (CES-D) | |
| Beauboeuf-Lafontant (2007) | To investigate the possible relationship between constructions of strong Black womanhood and the experiences of silencing and selflessness suggested in the clinical literature on women's experiences of depression. | US | Interviews | Grounded Theory, Voice centred | 44 | Black American | Mean: 35. Range: 19 to 67 | No | Not Stated |
| Beauboeuf-Lafontant (2008)(Beauboeuf-Lafontant, 2008) | To empirically examine both the construction of good Black womanhood as "being strong" and its possible contributions to depression. | US | Interviews | Voice centred | 58 | African American | Mean: 35.6. Range: 19 to 67 | Yes (self-disclosure) sought out a non-clinical sample | No |
| Besson (2012) | To understand the experiences of Black women receiving culturally sensitive care | US | Interviews | Interpretative phenomenological analysis | Total 8: Black women 6 | African American | Mean: 52 (SD, 5.49). Range: 41 to 57 | Yes (validated questionnaire) Quick Inventory of Depression | Yes |

| | | | | | | | | | |
|---|---|---|---|---|---|---|---|---|---|
| | within a cross-racial context. | | | | | | | Symptomatolog y | |
| Black (2007) | What is the meaning of depression to a group of elderly African American women? And how is it experienced and expressed? | US | Interviews (ethnographic ) and informal conversation | Thematic and Narrative analysis | Total 120: Back women 20 | African American | 80+ | Yes (self-disclosure) | Not Stated |
| Black (2011) | To understand the cultural traditions and religious/spiritual beliefs that underlie experiences and expressions of depression, and elders' perceived causes and preferred resolutions. | US | Interviews | Large-level sorting and fine-grain analysis | Total 60: Black women 30 | African American | Mean: 73.1 (SD, 6). Range: 60 to 88 | Yes (self-disclosure) | Not Stated |
| Borum (2012) | To examine perceptions of risk (e.g., depression) and protection (e.g., the role of spirituality) associated with suicide in the lives of 40 African American women. | US | Focus Groups | Grounded Theory Content Analysis | 40 | African American | Mean: 26. Range: 18 to 39 | Not Stated | Not Stated |

| | | | | | | | | | |
|---|---|---|---|---|---|---|---|---|---|
| Brown (2011) | To qualitatively examine reasons why women would decide not to consult their GP if they experienced depressive symptoms and whether there might be ethno-cultural differences in this decision. | UK | Open-ended questionnaire | Inductive analysis, which combines elements of grounded theory | Total 145: Black women 73 | Black African, Black Caribbean, Black other | Range: 18 to 45. Mean: 31.32 | Yes (self-disclosure) | Yes |
| Cadigan (2015) | To elicit and examine their own Explanatory Models - the beliefs and assumptions they held about depression or depressive symptoms and how these understandings may have guided their help-seeking behaviours. | US | Interviews (ethnographic ) | Content Analysis | Total 32: Black women 15 | African American | Range: 16 to 40 | No | Not Stated |
| Campbell (2014) | To look more closely at the impact of culture, specifically cultural beliefs, on help-seeking behaviour and service use for depression among black Americans. | US | Interviews | Thematic Analysis | Total 17: Black women 13 | African American | Range: 21 to 57 | Yes (questions developed by the author) “1. having felt sad, empty, or depressed for two weeks or | Not Stated |

| | | | | | | | | | |
|---|---|---|---|---|---|---|---|---|---|
| | | | | | | | | more during their life; 2. having been told by a doctor, pastor, coworker, family member, or friend that he or she was depressed; 3. having seen a doctor or mental health professional for depression". | |
| Campbell (2016) | To explore Black Americans' experiences with depression, including experiences of stigma in Black communities. | US | Interviews | Thematic Analysis | Total 17: Black women 13 | African American | Range: 21 to 57 | Yes (questions developed by the author): "1. having felt sad, empty, or depressed for two weeks or more during their life; 2. having been told by a doctor, | Not Stated |

| | | | | | | | | | |
|---|---|---|---|---|---|---|---|---|---|
| | | | | | | | | pastor, coworker, family member, or friend that he or she was depressed; 3. having seen a doctor or mental health professional for depression." | |
| Clements (2023) | How do African American women use social support networks to address depression and the SBW schema? | US | Interviews | Thematic Analysis | 16 | African American | Range: 25 to 50 | Yes (self-disclosure) | Not Stated |
| Conner (2010) | 1. To examine the experience of being depressed among African American elders and their perceptions of barriers confronted when contemplating seeking mental health services. 2. To examine how coping strategies are utilised by | US | Interviews | Thematic Analysis and Content Analysis | 37 | African American | Range: 60 to 81 | Yes (validated questionnaire) Patient Health Questionnaire (PHQ-9) a score of 5 or above | Not Stated |

| | | | | | | | | | |
|---|---|---|---|---|---|---|---|---|---|
| | African-American elders who choose not to seek professional mental health services. | | | | | | | | |
| Copeland (2011) | To explore why mothers who seek mental health services for their children do not receive mental health services for themselves | US | Interviews (ethnographic) | Thematic Analysis | 32 | African American | Mean: 37. Range: 23 to 62 | Yes (validated questionnaire) Patient Health Questionnaire and the Beck Depression and Anxiety Inventory. A score of 10 and above on the BDI and BAI or a positive score on the PHQ | Not Stated |
| Curtis (2018) | To learn more about how these parents, and in particular mothers, understand their experiences of psychological distress. | US | Interviews | Grounded Theory and Narrative Analysis | Total 13: Black women 5 | African American | Range: 23 to 50 | Yes (validated questionnaire) Either the MINI-MDE questionnaire or the PHQ-2 scale | Not Stated |

| | | | | | | | | | |
|---|---|---|---|---|---|---|---|---|---|
| Dare (2023) | How do University students of African, Caribbean and similar ethnicity conceptualise mental illness and help-seeking behaviours? | UK | Interviews | Thematic Analysis | Total 6: Black women 3 | Black African or Caribbean | Range: 22 to 55 | Yes (self-disclosure) | Not Stated |
| Desai (2023) | To study the local psychosocial pathways to depressive and significantly distressing experiences, specifically examining the role of racism and racialised structure. | US | Interviews and focus groups | Interpretative phenomenological analysis | 25 total. 17 were women, but it is unclear how many were Black women | Black or African American | Mean for focus group participants: 50. Mean for interview participants: 48 | No | Not Stated |
| Dunlap (2022) | What are the lived experiences of the depressed and/or anxious strong Black woman? | US | Interviews | Interpretative phenomenological analysis | 10 | African American | Range: 31 to 50 | Yes (nominated by a mental health professional) | Yes |
| Etowa (2007) | The study explored first-person experiences of health and well-being among midlife Nova Scotian women of African descent with a goal of gathering baseline qualitative and | Canada Nova Scotia | Interviews Focus Groups Community workshops to present findings and incorporate interpretation | Thematic Analysis | 113 | African Canadian | Mean: 51.78 (SD, 6.24). Range: 40 to 65 | Yes (validated questionnaire) Centre for Epidemiological Studies on Depression (CES-D) questionnaire | Not Stated |

| | | | | | | | | | |
|---|---|---|---|---|---|---|---|---|---|
| | quantitative data concerning their perceptions of depression general health status, menopause symptoms, race-related stress, and healthy lifestyle indicators. | | into the analysis | | | | | | |
| Graham (2021) | 1. How do Black women experience and manage symptoms of depression and anxiety? 2. To what extent does the SBW image shape and inform and provide a context for Black women's experiences and management of depressive and anxiety-related symptoms? | UK | Focus Groups | Thematic Analysis | 18 | African Caribbean | Mean: 42. Range: 19 to 57 | Yes (self-disclosure) | No |
| Holden (2015) | To foster a greater understanding of diverse African American women's perceptions about psychosocial, socio-cultural, and | US | Focus Groups | Thematic Analysis | 63 | African American | Mean: 45 (SD, 12.05). Range: 22 to 64 | Not Stated | Not Stated |

| | | | | | | | | | |
|---|---|---|---|---|---|---|---|---|---|
| | environmental related contributors to depression. | | | | | | | | |
| Hong (2023) | To examine the association of mindfulness, depression, and depression stigma among African American women who participated in an eight-week mindfulness-based depression intervention. | US | Focus Groups | Thematic Analysis | 24 | African American | Mean: 59.9 | Yes (validated questionnaire) Inventory of Depressive Symptomatolog y – Clinician Rated (IDS-C) and the Quick Inventory of Depressive Symptomatolog y (QIDS-C) | Yes |
| Keller (2016) | To explore the experience of depression disclosure in primary care settings among self-identified African American, Hispanic and non-Hispanic white women. | US | Interviews | Content Analysis (inductive) | Total 24: Black women 9 | African American | Mean: 40. Range: 18 to 58 | Yes (validated questionnaire) PHQ-8 scores ranged from 3 to 24 | Yes |

| | | | | | | | | | |
|---|---|---|---|---|---|---|---|---|---|
| Mitchell (2005) | 1. What is the range of definitions, thoughts, feelings, and views held by African American women relative to professional mental health treatment? 2. What are the perceived barriers and incentives to seeking out formal mental health care? 3. What are the kinds of problems that African American women consider appropriate for this form of help? | US | Interviews | Constant Comparative methods | 8 | African American | Range: 21 to 55 | Not Stated | Yes |
| Nelson (2020) | 1. How did Black women conceptualise help-seeking for depression? 2. How was the help-seeking process informed by the Strong Black Woman role? | US | Interviews | Thematic Analysis | 30 | African American, Caribbean (e.g., Barbados, Dominican Republic, Haiti, and Jamaica) | Mean: 33.43 (SD, 12.43). Range: 18 To 66 | Not Stated | Not Stated |

| | | | | | | | | | |
|---|---|---|---|---|---|---|---|---|---|
| Nicolaidis (2010) | To understand the experiences and beliefs of depressed African American women residing in Portland, Oregon, a city with relatively low racial diversity, regarding depression and depression care. | US | Focus Groups | Thematic Analysis | 30 | African American | Mean: 36.2. Range: 19 to 53 | Yes (validated questionnaire) PHQ9, score15 or higher | Yes |
| Progovac (2020) | To better understand (a) how past experiences of health care discrimination play a role in the experience of seeking and receiving mental health treatment, (b) whether and how treatment preferences are elicited by providers and then routinely incorporated into clinical care for depression, and (c) to what degree treatment preferences are shaped by experiences of | US | Interviews | Thematic Analysis | Total 21: Black women 8 | Non-Hispanic/Latino Black | Range: 18 to 60 | Yes (self-disclosure (validated questionnaire PHQ9) | Not Stated |

| | | | | | | | | | |
|---|---|---|---|---|---|---|---|---|---|
| | health care discrimination. | | | | | | | | |
| Schreiber (1998) | To explore the important contextual issues concerning black West Indian Canadian women's experiences with depression. | Canada | Interviews | Grounded Theory | 12 | West Indian | Mean: 39 | Yes (self-disclosure) | Not Stated |
| Schreiber (2000) | To discover how women from a non-dominant cultural background (West Indian) experience and manage depression. | Canada | Interviews and observations | Grounded Theory | 12 | West-Indian Canadian women | N/A | No | No |
| Sellers (2006) | What are the major health and well-being issues confronting African immigrant women residing in the USA? What dimensions are associated with these concerns? What type of interventions would | US | Focus Groups | Dimensional Analysis and Constant Comparative methods | 5 | Black or African American | Mean: 43.4. Range: 39-48 | No | No |

| | | | | | | | | | |
|---|---|---|---|---|---|---|---|---|---|
| | participants perceive as culturally appropriate treatment for their health and well-being concerns? | | | | | | | | |
| Sisley (2011) | To explore individual explanatory models of experiences of distress, coping and help-seeking choices to improve the cultural relevance of services. | UK | Interviews | Interpretative phenomenological analysis | 7 | African Caribbean | Range: 30 to 50 | Yes (self-disclosure) | Not Stated |
| Waite (2007) | a) How women came to understand that they were depressed, (b) words and language used to describe depression, (c) how women talked about depression in their everyday life, (d) how women labelled their feelings, and (e) how | US | Focus Groups | Content Analysis | 36 | African American, Black American | Range: 35 to 45 | Yes (validated questionnaire) PHQ 9 | Not Stated |

| | | | | | | | | | |
|---|---|---|---|---|---|---|---|---|---|
| | women constructed meaning from their personal experience for dealing with depression. | | | | | | | | |
| Waite (2008) | To explore and describe the health beliefs of African American women about depression to improve insight regarding treatment decisions. | US | Focus Groups | Thematic Analysis | 14 | African American | Mean: 45 (SD, 10·50). Range: 18 to 64 | Yes (self-disclosure and PHQ9) | Yes |
| Waite (2009) | 1. To investigate the core components of explanatory models about depression among a cohort of low-income African American women. 2. To examine how explanatory models shape a person's response towards seeking help for depression. | US | Focus Groups | Content Analysis and Constant Comparative methods | 14 | African American | Mean: 40. Range: 25 to 65 | Yes, participants had to be diagnosed with Major Depressive Disorder by a mental health provider within the past year. | Yes |

| | | | | | | | | | |
|---|---|---|---|---|---|---|---|---|---|
| Walton (2019) | The study aimed to explore experiences of depression relevant to the intersections of race, class, and gender for Black women with middle-class SES. We examined how contextual factors (i.e., personal, social, and cultural) shape how depression is perceived. | US | Interviews | Thematic Analysis | 30 | Black | Mean: 37.66 (SD, 4.54). Range: 30 to 45 | Yes (validated questionnaire) PHQ9 | Not Stated |
| Walton (2021) | To explore how the identity of being a middle-class Black woman informed their experiences of depression. | US | Interviews | Grounded Theory and Constant Comparative methods | 30 | Black | Mean: 37.66 (SD, 4.54). Range: 30 to 45 | Yes (validated questionnaire) PHQ9 | Not Stated |
| Ward (2014) | Examined older African American women's lived experiences with depression and coping behaviours. | US | Interviews | Interpretative phenomenological analysis | 13 | African American | Mean: 71. Range: 60 to 78 | Yes (validated questionnaire) Center for Epidemiologic Studies Depression Scale, mean | Not Stated |

| | | | | | | | | | |
|---|---|---|---|---|---|---|---|---|---|
| | | | | | | | | CES-D score of 24.5 | |
| Wharton (2018) | Examined qualitative data from older, church-going African Americans to explore issues relevant to their attitudes and expectations around formal mental health care. | US | Focus Groups | Constant Comparative methods | Total 50: Black women 29 | African American | Mean: 67. Range: 51 to 94 | No | No |
| Wittink (2009) | To gain insight into the process of help-seeking and the identification and treatment of depression among older African Americans in primary care. | US | Interviews | Constant Comparative methods | Total 47: Black women 37 | African American | Mean: 783 | Yes (validated questionnaire), Centres for Epidemiologic Studies Depression scale | Not Stated |

##### Appendix E: Critical Appraisal Skills Programme (CASP) results

| Author | Was there a clear statement of the aims of the research? | Is a qualitative methodology appropriate? | Was the research design appropriate to address the aims of the research? | Was the recruitment strategy appropriate to the aims of the research? | Was the data collected in a way that addressed the research issue? | Has the relationship between researcher and participants been adequately considered? | Have ethical issues been taken into consideration? | Was the data analysis sufficiently rigorous? | Is there a clear statement of findings? | Does the researcher discuss the contribution the study makes to existing knowledge or understanding (e.g. do they consider the findings in relation to current practice or policy or relevant research-based literature) |
|---|---|---|---|---|---|---|---|---|---|---|
| Akinyemi (2018) | Yes | Yes | Yes | Yes | Yes | No | Yes | Yes | Yes | Yes |
| Alang (2016) | Yes | Yes | Yes | Yes | Yes | Yes | Yes | Yes | Yes | Yes |
| Anderson (2021) | Yes | Yes | Yes | Yes | Yes | Yes | Yes | Yes | Yes | Yes |

| | | | | | | | | | | |
|---|---|---|---|---|---|---|---|---|---|---|
| Atanmo-Strempek (2014) | Yes | Yes | Yes | Yes | Yes | Yes | Yes | Yes | Yes | Yes |
| Bailey (2021) | Yes | Yes | Yes | Yes | Yes | No | Can't Tell | Yes | Yes | Yes |
| Barbee (1994) | Yes | Yes | Yes | Yes | Can't Tell | No | Can't Tell | Yes | Yes | Yes |
| Beagan (2012) | Yes | Yes | Yes | Yes | Yes | Yes | Yes | Yes | Yes | Yes |
| Beauboeuf-Lafontant (2007) | Yes | Yes | Yes | Yes | Yes | Yes | Can't Tell | Yes | Yes | Yes |
| Beauboeuf-Lafontant (2008) | Yes | Yes | Yes | Yes | Yes | Yes | Can't Tell | Yes | Yes | Yes |
| Besson (2012) | Yes | Yes | Yes | Yes | Yes | Yes | Yes | Yes | Yes | Yes |
| Black (2007) | Yes | Yes | Yes | Yes | Yes | No | Yes | Yes | Yes | Yes |
| Black (2011) | Yes | Yes | Yes | Yes | Yes | Yes | Yes | Yes | Yes | Yes |
| Borum (2012) | Yes | Yes | Yes | Yes | Yes | Yes | Yes | Yes | Yes | Yes |
| Brown (2011) | Yes | Yes | Yes | Yes | Yes | Yes | Yes | Yes | Yes | Yes |
| Cadigan (2015) | Yes | Yes | Yes | Yes | Yes | Yes | Yes | Yes | Yes | Yes |
| Campbell (2014) | Yes | Yes | Yes | Yes | Yes | Yes | Yes | Yes | Yes | Yes |
| Campbell (2016) | Yes | Yes | Yes | Yes | Yes | Yes | Can't Tell | Yes | Yes | Yes |
| Clements (2023) | Yes | Yes | Yes | Yes | Yes | Yes | Yes | Yes | Yes | Yes |

| | | | | | | | | | | |
|---|---|---|---|---|---|---|---|---|---|---|
| Conner (2010) | Yes | Yes | Yes | Yes | Yes | No | Yes | Yes | Yes | Yes |
| Copeland (2011) | Yes | Yes | Yes | Yes | Yes | Yes | Yes | Yes | Yes | Yes |
| Curtis (2018) | Yes | Yes | Yes | Yes | Yes | Yes | Yes | Yes | Yes | Yes |
| Dare (2023) | Yes | Yes | Yes | Yes | Yes | Yes | Yes | Yes | Yes | Yes |
| Desai (2023) | Yes | Yes | Yes | Yes | Yes | Yes | Yes | Yes | Yes | Yes |
| Dunlap (2022) | Yes | Yes | Yes | Yes | Yes | Yes | Yes | Yes | Yes | Yes |
| Etowa (2007) | Yes | Yes | Yes | Yes | Yes | Yes | Yes | Yes | Yes | Yes |
| Graham (2021) | Yes | Yes | Yes | Yes | Yes | Yes | Yes | Yes | Yes | Yes |
| Holden (2015) | Yes | Yes | Can't Tell | Yes | Yes | Yes | Yes | Yes | Yes | Yes |
| Hong (2023)(Hong, Satyshur, & Burnett-Zeigler, 2023) | Yes | Yes | Yes | Yes | Yes | Yes | Yes | Yes | Yes | Yes |
| Keller (2016)(Keller, Valdez, Schwei, & Jacobs, 2016) | Yes | Yes | Yes | Yes | Yes | No | Yes | Yes | Yes | Yes |
| Mitchell (2005)(Mitchell, 2005) | Yes | Yes | Yes | Yes | Yes | Yes | Yes | Yes | Yes | Yes |

| | | | | | | | | | | |
|---|---|---|---|---|---|---|---|---|---|---|
| Nelson (2020)(Nelson, Shahid, & Cardemil, 2020) | Yes | Yes | Yes | Yes | Yes | Yes | Yes | Yes | Yes | Yes |
| Nicolaidis (2010)(Nicolaidis , et al., 2010) | Yes | Yes | Yes | Yes | Yes | Yes | Yes | Yes | Yes | Yes |
| Progovac (2020)(Progovac, et al., 2020) | Yes | Yes | Yes | Yes | Yes | Yes | Yes | Yes | Yes | Yes |
| Schreiber (1998)(Rita Schreiber, Stern, & Wilson, 1998) | Yes | Yes | Yes | Yes | Yes | Yes | Yes | Yes | Yes | Yes |
| Schreiber (2000)(R. Schreiber, Stern, & Wilson, 2000) | Yes | Yes | Yes | Can't Tell | Yes | Yes | Can't Tell | Yes | Yes | Yes |
| Sellers (2006)(Sellers, Ward, & Pate, 2006) | Yes | Yes | Can't Tell | Yes | Yes | Yes | Yes | Yes | Yes | Yes |
| Sisley (2011)(Sisley, Hutton, Louise | Yes | Yes | Yes | Yes | Yes | No | Yes | Yes | Yes | Yes |

| | | | | | | | | | | |
|---|---|---|---|---|---|---|---|---|---|---|
| Goodbody, & Brown, 2011) | | | | | | | | | | |
| Waite (2007)(Roberta Waite & Killian, 2007) | Yes | Yes | Can't Tell | Yes | Yes | Yes | Yes | Yes | Yes | Yes |
| Waite (2008) | Yes | Yes | Yes | Yes | Yes | Yes | Yes | Yes | Yes | Yes |
| Waite (2009) | Yes | Yes | Yes | Yes | Yes | Can't Tell | Yes | Yes | Yes | Yes |
| Walton (2019) | Yes | Yes | Yes | Yes | Yes | Yes | Yes | Yes | Yes | Yes |
| Walton (2021) | Yes | Yes | Yes | Yes | Yes | Yes | Yes | Yes | Yes | Yes |
| Ward (2014) | Yes | Yes | Yes | Yes | Yes | Yes | Yes | Yes | Yes | Yes |
| Wharton (2018) | Yes | Yes | Yes | Yes | Yes | Yes | Yes | Yes | Yes | Yes |
| Wittink (2009) | Yes | Yes | Yes | Yes | Yes | Can't Tell | Yes | Yes | Yes | Yes |

#### Appendix F: Detailed Confidence in the Evidence from Reviews of Qualitative Research (GRADE-CERQual) Assessments

| Summary of review finding | Studies contributing to the review finding | Relevance | Methodological Limitations | Coherence | Adequacy | GRADE-CERQual assessment of confidence in the evidence | Explanation of GRADE-CERQual assessment |
|---|---|---|---|---|---|---|---|
| 1. **Depression stemming from adverse life experiences** This finding suggested that various types of adverse life experiences were associated with Black women's experiences of depression. The types of adverse experiences were varied (poverty, different types of abuse and neglect from Black men). Racial trauma was cumulative, leading to racial battle fatigue and mental health breakdowns | (Anderson, 2021; Beagan, Etowa, & Bernard, 2012; Beauboeuf-Lafontant, 2007, 2008; H. K. Black, White, & Hannum, 2007; Copeland & Snyder, 2011; Curtis, Morgan, & Laird, 2018; Dunlap, 2022; Graham & Clarke, 2021; Holden, Belton, & Hall, 2015; Nelson, et al., | No concerns. Only one(Curtis, et al., 2018) study included a mixed sample. The study included other ethnicities, but we only synthesised data from Black women. | Very minor concerns. Two studies did not mention the position of the researcher.(H. K. Black, et al., 2007; Sisley, et al., 2011) Two other studies did not report whether they had secured ethical approval.(Beauboeuf-Lafontant, 2007, 2008) | No concerns. The finding reflects the variations in the adverse experience that might lead to depression, it seemed supported with the detail provided. | No Concerns. Theme was both descriptive and explanatory and was supported by multiple studies, which included both thin data (for descriptive) and rich data (for explanatory). | High confidence | Seventeen papers contributed to this theme, and there were very minor or no concerns across all categories. |

| | | | | | | | |
|---|---|---|---|---|---|---|---|
| | 2020; Nicolaidis, et al., 2010; Rita Schreiber, et al., 1998; Sisley, et al., 2011; Roberta Waite & Killian, 2007; R. Waite & Killian, 2009; Ward, Mengesha, & Issa, 2014) | | | | | | |
| **1.1 Mistrust of clinicians resulting in reluctance towards pharmacological interventions** This finding suggested that Black women did not trust healthcare providers. The lack of trust was associated with fear. The fear stemmed from being treated poorly, facing judgement about their ability | (Alang, 2016; Bailey & Tribe, 2021; Beagan, et al., 2012; Cadigan & Skinner, 2015; Campbell & Long, 2014; Campbell & Mowbray, 2016; Conner, et al., 2010; Copeland & | Moderate concerns. Eight of the included studies(Alang, 2016; Bailey & Tribe, 2021; Cadigan & Skinner, 2015; Campbell & Long, 2014; Campbell & Mowbray, 2016; Keller, et al., 2016; Progovac, et | Minor concerns. Four studies (Bailey & Tribe, 2021; Conner, et al., 2010; Keller, et al., 2016; Sisley, et al., 2011) did not mention the position of the researcher and it was unclear in one.(R. Waite & Killian, 2009) In two studies it was unclear if | No concern. The interpretation seems to be supported by the data. The data shows the various interpretations (types of fear etc.) and organises them into a | No concerns. Theme was both descriptive and explanatory and was supported by multiple studies, which included both thin data (for | High confidence | Nineteen papers contributed to this theme, and there were minor or no concerns across all categories except for relevance, which was moderate. This did not affect our confidence in the findings. |

| | | | | | | | |
|---|---|---|---|---|---|---|---|
| to take care of themselves and their loved ones, concerns about breach in confidentiality and experiencing discrimination from White clinicians. Consequently, women were reluctant to accept pharmacological interventions. Women were open to talk therapies. Depending on the rapport they built with their therapist, women continued to utilise the service and, in some cases, could be convinced to take medication alongside therapy. The rapport between the patient and therapist was vital. | Snyder, 2011; Holden, et al., 2015; Keller, et al., 2016; Mitchell, 2005; Nelson, et al., 2020; Nicolaidis, et al., 2010; Progovac, et al., 2020; Sellers, et al., 2006; Sisley, et al., 2011; Roberta Waite & Killia, 2008; R. Waite & Killian, 2009; Wharton, Watkins, Mitchell, & Kales, 2018) | al., 2020; Wharton, et al., 2018) had mixed sample, but we only synthesised data from Black women. | ethical approval was secured.(Bailey & Tribe, 2021; Campbell & Mowbray, 2016) | narrative that shows various sides. | descriptive) and rich data (for explanatory). For instance, the reasons for the lack of trust may not have a lot of studies for each point but included very rich data which we could derive explanatory findings from. | | |
| **1.2 Access to a representative, compassionate and effective clinician** | (Bailey & Tribe, 2021; Besson, 2012; Helen K. Black, Gitlin, & Burke, 2011; | Minor concerns. Six studies(Bailey & Tribe, 2021; Helen K. Black, et al., 2011; | Minor concerns. It was unclear if ethics was considered in two studies.(Bailey & Tribe, 2021; R. | No concerns. The data were varied and used to present different | No Concerns. Theme explored the type of therapist | High confidence | Twenty papers contributed to this theme, and there were minor or no |

| | | | | | | | |
|---|---|---|---|---|---|---|---|
| This finding suggested that women preferred clinicians who were relatable, either shared identity or had experience working with Black women. This theme suggested that visible characteristics or perceived understanding of their lived experiences initiated service utilisation. However, retention was only maintained if the psychotherapist was compassionate and effective. | Campbell & Long, 2014; Copeland & Snyder, 2011; Dare, Jidong, & Premkumar, 2023; Graham & Clarke, 2021; Keller, et al., 2016; Mitchell, 2005; Nelson, et al., 2020; Nicolaidis, et al., 2010; Progovac, et al., 2020; R. Schreiber, et al., 2000; Sellers, et al., 2006; Sisley, et al., 2011; Roberta Waite & Killia, 2008; R. Waite & Killian, 2009; Ward, et al., 2014; Wharton, | Campbell & Long, 2014; Keller, et al., 2016; Progovac, et al., 2020; Wharton, et al., 2018; Wittink, et al., 2009) included a mixed sample, but we only synthesis data from Black women. | Schreiber, et al., 2000) The position of author was not reported in three studies (Bailey & Tribe, 2021; Keller, et al., 2016; Sisley, et al., 2011), and it was unclear for two studies.(R. Waite & Killian, 2009; Wittink, et al., 2009) There were other components not well reported such as research design and recruitment strategy, but they were unlikely to affect our confidence in the QES. | expectations and reasons behind for the type of therapist they wanted to work with. | women wanted to work with and there was both descriptive and explanatory reasons which were supported by multiple studies, with both thin data (for descriptive) and rich data (for explanatory). For instance, one quote describes why they needed a clinician that looked like them. It was | | concerns across all categories. |

| | | | | | | | |
|---|---|---|---|---|---|---|---|
| | et al., 2018; Wittink, Joo, Lewis, & Barg, 2009) | | | | rich enough for an explanatory theme. | | |
| **2. Social Isolation** This theme suggested that women had periods of wanting to be alone and feeling lonely. Wanting to be alone was driven by a need for social distance to recalibrate or hide from the stigma associated with depression. At times, women disclosed that they were depressed by their social network when they did not understand their experiences. This was associated with women feeling stigmatised for being depressed, so they withdrew. | (Atanmo-Strempek, 2014; Barbee, 1994; Beagan, et al., 2012; Helen K. Black, et al., 2011; H. K. Black, et al., 2007; Borum, 2012; Clements, 2023; Conner, et al., 2010; Graham & Clarke, 2021; Nelson, et al., 2020; Rita Schreiber, et al., 1998; R. Schreiber, et al., 2000; Sellers, et al., 2006; | No concerns. One study(Helen K. Black, et al., 2011) included a mixed sample, but we only synthesised data from Black women. | Minor concerns. Ethical considerations were unclear in two studies.(Barbee, 1994; R. Schreiber, et al., 2000) The position of the author was not mentioned in four studies(Barbee, 1994; H. K. Black, et al., 2007; Conner, et al., 2010; Sisley, et al., 2011) and it was unclear in one.(R. Waite & Killian, 2009) A couple of other studies did not report their recruitment strategy, and the research | Minor concerns. The data shows that women wanted to be alone, or they felt lonely. However, the reasons for how they felt is not supported by multiple studies. There is a strong on description but not very strong on the explanatory parts of the finding. | Minor concerns. The main finding was that women wanted to be alone, and they also felt lonely. This was supported by data from several studies. However, the explanatory section on why they wanted for be alone e.g., was not | High confidence | Seventeen papers contributed to this theme, and there were very minor or no concerns across all categories. |

| | | | | | | | |
|---|---|---|---|---|---|---|---|
| | Sisley, et al., 2011; Roberta Waite & Killian, 2007; R. Waite & Killian, 2009; Ward, et al., 2014) | | design was unclear, but they were unlikely to affect our confidence in the QES. | | supported by multiple studies. | | |
| **2.1 Stigma surrounding depression** This theme suggested that women experienced stigma generally associated with depression and stigma specific to the Black community. Women feared they would be considered “crazy”, so women only disclosed their experiences of depression to people they trusted, which included family and friends. Others only shared with family as there was cultural belief that it should only be kept in the family. The stigma resulted in | (Alang, 2016; Atanmo-Strempek, 2014; Bailey & Tribe, 2021; Barbee, 1994; Beagan, et al., 2012; Beauboeuf-Lafontant, 2007; Besson, 2012; Helen K. Black, et al., 2011; H. K. Black, et al., 2007; Borum, 2012; Campbell & Long, 2014; Campbell & | Very minor. Eight studies(Alang, 2016; Bailey & Tribe, 2021; Helen K. Black, et al., 2011; Campbell & Long, 2014; Campbell & Mowbray, 2016; Curtis, et al., 2018; Dare, et al., 2023; Desai, et al., 2023) included a mixed sample, but we only synthesised data from Black women. | Moderate concern. Ethical considerations were not clear six studies.(Bailey & Tribe, 2021; Barbee, 1994; Beauboeuf-Lafontant, 2007, 2008) (Campbell & Mowbray, 2016; R. Schreiber, et al., 2000) The position of the author was not considered in the seven studies (Akinyemi, et al., 2018; Bailey & Tribe, 2021; Barbee, | No concerns. The data were varied and used to present different types of and reasons behind the type of coping women utilised. There is a good mix of descriptive and interpretation of findings. | No Concerns. Theme explored the different types of stigmas, where the stigma stemmed from and how it affected Black women's help-seeking behaviours. Each point was supported by | High confidence | Thirty-five papers contributed to this theme, and there were no concerns or very minor concerns across all categories except for methodological limitations, which did not affect our confidence in the findings. |

| | | | | | |
|---|---|---|---|---|---|
| women needing to tackle the experience of depression on their own, which fed into social isolation. | Mowbray, 2016; Clements, 2023; Conner, et al., 2010; Copeland & Snyder, 2011; Curtis, et al., 2018; Dare, et al., 2023; Desai, et al., 2023; Etowa, Keddy, Egbeyemi, & Eghan, 2007; Graham & Clarke, 2021; Holden, et al., 2015; Hong, et al., 2023; Mitchell, 2005; Nelson, et al., 2020; Nicolaidis, et al., 2010; Rita Schreiber, et al., 1998; R. Schreiber, et al., | | 1994; H. K. Black, et al., 2007; Conner, et al., 2010; Keller, et al., 2016; Sisley, et al., 2011) and it was unclear on two studies.(R. Waite & Killian, 2009; Wittink, et al., 2009) A couple other studies did not report their recruitment strategy and research design but this did not affect our confidence QES. | | several studies. Data were both descriptive and explanatory reasons which were supported by multiple studies, with both thin data (for descriptive) and rich data (for explanatory). For instance, one quote describes that there is a different type of stigma associated with being Black female |

| | | | | | | | |
|---|---|---|---|---|---|---|---|
| | 2000; Sellers, et al., 2006; Sisley, et al., 2011; Roberta Waite & Killia, 2008; Roberta Waite & Killian, 2007; R. Waite & Killian, 2009; Walton, 2021; Walton & Boone, 2019; Ward, et al., 2014) | | | | and middle class. It was rich enough for an explanatory theme. | | |
| **3. Navigating depression within a framework of faith** This theme suggested that religion was considered historically and culturally embedded in the black community. Therefore, women who experienced depression as a loss of faith felt ashamed | (Akinyemi, et al., 2018; Atanmo-Strempek, 2014; Bailey & Tribe, 2021; H. K. Black, et al., 2007; Cadigan & Skinner, 2015; Campbell & Long, 2014; | Very minor concerns. Five studies(Akinyemi, et al., 2018; Bailey & Tribe, 2021; Cadigan & Skinner, 2015; Campbell & Long, 2014; Wittink, et al., 2009) included a mixed sample, | Minor concerns. Ethical considerations were unclear in 1 study.(Bailey & Tribe, 2021) The position of the author not mentioned in two studies(Akinyemi, et al., 2018; H. K. Black, et al., 2007) | Minor concerns. The data shows that there was a spiritual element to how Black women experienced depression. However, the | Very minor concerns. The main finding was that women experienced depression as a lack of faith. This was supported by | High confidence | Thirteen papers contributed to this theme, and there were very minor or minor concerns across all categories. |

| | | | | | | | |
|---|---|---|---|---|---|---|---|
| | Etowa, et al., 2007; Mitchell, 2005; Roberta Waite & Killia, 2008; R. Waite & Killian, 2009; Walton, 2021; Walton & Boone, 2019; Wittink, et al., 2009) | but we only synthesised data from Black women. | and it was unclear in two other studies. (R. Waite & Killian, 2009; Wittink, et al., 2009) | interpretation on feeling ashamed is really only mentioned in one study. Not a great concern as the main narrative is that Black there a is faith element associated with depression. | data from several studies. So was the explanation regarding why religion plays a pivotal role. However, the explanatory section that it made them feel ashamed was only reported in one study. | | |
| **3.1 Religious coping** This theme suggested that women utilised religious coping to manage episodes of depression. Religious coping included prayer, reading the Bible, speaking to their faith leader, and praise and | (Anderson, 2021; Atanmo-Strempek, 2014; Bailey & Tribe, 2021; Barbee, 1994; Beagan, et al., 2012; Beauboeuf- | Minor concerns. Seven studies(Bailey & Tribe, 2021; Helen K. Black, et al., 2011; Brown, et al., 2011; Cadigan & | Minor concerns. Ethical considerations unclear in five studies.(Bailey & Tribe, 2021; Barbee, 1994; Beauboeuf-Lafontant, 2007, | No concerns. The data were varied and used to present different types of religious coping. There is a good mix | Very minor concerns. One of the findings was descriptive (different type of religious | High confidence | Thirty papers contributed to this theme, and there were very minor/no concerns or minor concerns across all categories. |

| | | | | | |
|---|---|---|---|---|---|
| worship. This prevented women from utilising formal mental health services as they had faith that they would recover, as God did not give them more than they could handle. | Lafontant, 2007, 2008; Besson, 2012; Helen K. Black, et al., 2011; H. K. Black, et al., 2007; Borum, 2012; Brown, et al., 2011; Cadigan & Skinner, 2015; Campbell & Long, 2014; Clements, 2023; Conner, et al., 2010; Curtis, et al., 2018; Etowa, et al., 2007; Graham & Clarke, 2021; Mitchell, 2005; Nelson, et al., 2020; Rita Schreiber, et al., 1998; R. Schreiber, et al., | Skinner, 2015; Campbell & Long, 2014; Curtis, et al., 2018; Wittink, et al., 2009) included a mixed sample, but we only synthesised data from Black women. | 2008; R. Schreiber, et al., 2000) The position of researcher not reported in four studies(Barbee, 1994; H. K. Black, et al., 2007; Conner, et al., 2010; Sisley, et al., 2011) and it was also unclear in two others.(R. Waite & Killian, 2009; Wittink, et al., 2009) A couple other studies did not report their recruitment strategy and research design, but this did not affect our confidence QES. | of descriptive and interpretation of findings. The explanatory findings having faith that they will be okay was supported by a lot of data and there was no data showing an alternative interpretation. | coping) and it was supported by several studies. Another complex finding on why they found solace in religious coping was supported by several rich studies. However, on last point on using it in response to oppression was only supported by one study. |

| | | | | | | | |
|---|---|---|---|---|---|---|---|
| | 2000; Sisley, et al., 2011; Roberta Waite & Killia, 2008; Roberta Waite & Killian, 2007; R. Waite & Killian, 2009; Walton, 2021; Ward, et al., 2014; Wittink, et al., 2009) | | | | | | |
| **3.2 Endorsement and support from faith leaders** This theme suggested that some women needed approval from faith leaders to utilise mental health services. Given the importance religion holds in the Black community, perhaps approval from faith leaders served as approval from the Black community, which may have reduced stigma. Once they accessed | (Roberta Waite & Killia, 2008; Wharton, et al., 2018; Wittink, et al., 2009) | Moderate concerns. Two of the three studies(Wharton, et al., 2018; Wittink, et al., 2009) included a mixed sample, but we only synthesised data from Black women. | No concern. It was unclear if the position of the researcher was considered in one study.(Wittink, et al., 2009) | Serious concerns. The interpretation of the theme is supported mainly supported by the richness of 1 study. | Moderate concern.<br><br>Very few studies although one of them is relatively rich and it shows a key part of the discussion. | Low confidence | Three papers contributed to this theme, and although there were no concerns about methodological limitations, there were moderate concerns about adequacy and relevance and serious concerns about coherence. |

| | | | | | | | |
|---|---|---|---|---|---|---|---|
| services, they realised they did not have to renounce their faith as they believed God worked through medicine. | | | | | | | |
| 4. **The Strong Black Woman schema masking depression symptoms**<br>This theme suggested that depression is a multifaceted phenomenon influenced by social and cultural factors in its manifestation. Many initially struggled to recognise symptoms as signs of depression. When they were not able to conform to the expectation of being a SBW and their functionality was compromised, they realised they were depressed. | (Akinyemi, et al., 2018; Alang, 2016; Atanmo-Strempek, 2014; Barbee, 1994; Beagan, et al., 2012; Beauboeuf-Lafontant, 2007; Helen K. Black, et al., 2011; H. K. Black, et al., 2007; Borum, 2012; Brown, et al., 2011; Cadigan & Skinner, 2015; Clements, 2023; Copeland & Snyder, 2011; | Very minor concerns. Six studies(Akinyemi, et al., 2018; Helen K. Black, et al., 2011; Brown, et al., 2011; Cadigan & Skinner, 2015; Curtis, et al., 2018; Desai, et al., 2023) included a mixed sample, but we only synthesised data from Black women. | Minor concerns. Ethical considerations were unclear in one study.(Beauboeuf-Lafontant, 2007) The position of the author was not reported in two studies.(H. K. Black, et al., 2007; Sisley, et al., 2011) | No concerns. The data shows that women were experiencing depression as weakness, and the different types of weaknesses and how it masked them. This show that different explanations were considered and reported. | Minor concerns. Fewer studies, but the data was highly relevant. The data shows that women were experiencing depression as weakness, but the different types of weaknesses were not supported by multiple studies, but they were rich. | High confidence | Thirty papers contributed to this theme, and there were no concerns, very minor or minor, across all categories. |

| | |
|---|---|
| | Curtis, et al., 2018; Desai, et al., 2023; Dunlap, 2022; Etowa, et al., 2007; Graham & Clarke, 2021; Holden, et al., 2015; Hong, et al., 2023; Mitchell, 2005; Nicolaidis, et al., 2010; R. Schreiber, et al., 2000; Sellers, et al., 2006; Sisley, et al., 2011; Roberta Waite & Killia, 2008; Roberta Waite & Killian, 2007; R. Waite & Killian, 2009; Walton & Boone, 2019; |

| | | | | | | | |
|---|---|---|---|---|---|---|---|
| | Ward, et al., 2014) | | | | | | |
| **4.1 Pressure to conform to being a Strong Black woman** This theme suggested that when Black women start to struggle to remain unphased by adversity, they try to prove that they are still strong Black women by denying that they are depressed. | (Alang, 2016; Bailey & Tribe, 2021; Barbee, 1994; Beagan, et al., 2012; Beauboeuf-Lafontant, 2007, 2008; Helen K. Black, et al., 2011; H. K. Black, et al., 2007; Borum, 2012; Cadigan & Skinner, 2015; Campbell & Long, 2014; Campbell & Mowbray, 2016; Clements, 2023; Conner, et al., 2010; Copeland & Snyder, 2011; Curtis, et al., | Very minor concerns. Seven studies(Bailey & Tribe, 2021; Helen K. Black, et al., 2011; Cadigan & Skinner, 2015; Campbell & Long, 2014; Campbell & Mowbray, 2016; Curtis, et al., 2018; Keller, et al., 2016) included a mixed but, we only synthesised data from Black women. | Moderate concern. Ethical considerations were unclear in the six studies.(Bailey & Tribe, 2021; Barbee, 1994) (Beauboeuf-Lafontant, 2007, 2008; Campbell & Mowbray, 2016; R. Schreiber, et al., 2000) The position of the research was not reported in six studies(Bailey & Tribe, 2021; Barbee, 1994; H. K. Black, et al., 2007; Conner, et al., 2010; Keller, et al., 2016; Sisley, et al., 2011) and it was unclear in one.(R. Waite & Killian, 2009) A couple other | Minor concerns. The data were varied and used to present different ways black women live up to being a SBW. There is a good mix of descriptive and interpretation of findings. | No concerns. There were several studies which both thin and rich data for each description and interpretation of the theme. | High confidence | Thirty-three papers contributed to this theme, and there were no concerns, very minor or minor, across all categories except for methodological limitations, which did not affect our confidence in the findings. |

| | | | |
|---|---|---|---|
| | 2018; Dunlap, 2022; Etowa, et al., 2007; Graham & Clarke, 2021; Holden, et al., 2015; Keller, et al., 2016; Mitchell, 2005; Nelson, et al., 2020; Nicolaidis, et al., 2010; Rita Schreiber, et al., 1998; R. Schreiber, et al., 2000; Sisley, et al., 2011; Roberta Waite & Killia, 2008; Roberta Waite & Killian, 2007; R. Waite & Killian, 2009; Walton, 2021; Walton & | | studies did not report their recruitment strategy and research design, but this did not affect our confidence QES. |

| | | | | | | | |
|---|---|---|---|---|---|---|---|
| | Boone, 2019; Ward, et al., 2014) | | | | | | |
| **4.2 Hitting a point of crisis leading to understanding depression** This theme suggested that women try to continue with their responsibilities, putting others before themselves until they hit a crisis point. The crisis is the wake-up call they need to take care of themselves and understand depression. Furthermore, what defines depression varies between populations depending on circumstances. | (Alang, 2016; Bailey & Tribe, 2021; Barbee, 1994; Beagan, et al., 2012; Beauboeuf-Lafontant, 2007; Helen K. Black, et al., 2011; H. K. Black, et al., 2007; Borum, 2012; Campbell & Long, 2014; Campbell & Mowbray, 2016; Clements, 2023; Conner, et al., 2010; Graham & Clarke, 2021; Holden, et al., 2015; Mitchell, | Very minor concerns. Six studies(Alang, 2016; Bailey & Tribe, 2021; Helen K. Black, et al., 2011; Campbell & Long, 2014; Campbell & Mowbray, 2016; Wharton, et al., 2018) included a mixed sample, but we only synthesised data from Black women. | Moderate concern. Ethical considerations were unclear in five studies.(Bailey & Tribe, 2021; Barbee, 1994; Beauboeuf-Lafontant, 2007; Campbell & Mowbray, 2016; R. Schreiber, et al., 2000) The position of the research was not reported in six studies(Bailey & Tribe, 2021; Barbee, 1994; H. K. Black, et al., 2007; Conner, et al., 2010; Keller, et al., 2016; Sisley, et al., 2011) and was unclear for one.(R. Waite & Killian, | No concerns. The data were varied and used to present different ways Black women understand depression The theme is quite descriptive. | No concerns. There were several studies which both thin and rich data for each description and interpretation of the theme. | High confidence | Twenty-seven papers contributed to this theme, and there were no concerns, very minor or minor, across all categories except for methodological limitations, which did not affect our confidence in the findings. |

| | | | |
|---|---|---|---|
| | 2005; Nelson, et al., 2020; Nicolaidis, et al., 2010; Rita Schreiber, et al., 1998; R. Schreiber, et al., 2000; Sisley, et al., 2011; Roberta Waite & Killia, 2008; Roberta Waite & Killian, 2007; R. Waite & Killian, 2009; Walton, 2021; Walton & Boone, 2019; Ward, et al., 2014; Wharton, et al., 2018) | | 2009) A couple other studies did not report their recruitment strategy and research design, but this did not affect our confidence QES. |
